# Seasonal and daily variation in indoor light and temperature associate with sleep disturbance in dementia

**DOI:** 10.1101/2024.10.30.24316428

**Authors:** Anne C Skeldon, Thalia Rodriguez Garcia, Eyal Soreq, Chloe Walsh, Derk-Jan Dijk, the UK Dementia Research Institute Care Research & Technology Centre

**Affiliations:** UK Dementia Research Institute Care Research & Technology Centre, Imperial College London, London and University of Surrey, Guildford, United Kingdom; School of Mathematics & Physics, Faculty of Engineering and Physical Sciences, University of Surrey, Guildford, United Kingdom; Surrey Sleep Research Centre, School of Biosciences, Faculty of Health and Medical Sciences, University of Surrey, Guildford, United Kingdom; Department of Brain Sciences, Imperial College London, United Kingdom; Surrey and Borders Partnership NHS Foundation Trust

## Abstract

Mechanisms regulating human sleep and physiology have evolved in response to rhythmic variation in environmental variables driven by the Earth’s rotation around its axis and the Sun. To what extent these mechanisms are operable in vulnerable people who are primarily exposed to the indoor environment remains unknown. We analysed 26,523 days of data from outdoor and indoor environmental sensors and a contactless behaviour-and-physiology sensor tracking bed occupancy, heart and breathing rate in 70 people living with dementia (PLWD). Indoor light and temperature, sleep timing, duration and fragmentation as well as the timing of the heart rate minimum all varied across seasons. Beyond the effects of season, higher bedroom temperature and less bright indoor daytime light associated with more disrupted sleep and higher respiratory rate. This sensitivity of sleep and physiology to ecologically relevant variations in indoor environmental variables implies that implementing approaches to control indoor light and temperature can improve sleep.

## Introduction

Sleep and circadian rhythmicity are important contributors to physical and mental health (Cederroth et al. 2019; Meyer et al. 2024). Sleep disturbances are predictors of, and prevalent in, neurodegenerative disorders (Wang and Holtzman 2020). The characteristics and intensity of these disturbances vary within and across disorders (Falgas et al. 2023; Shen et al. 2023; Benca and Teodorescu 2019; Koren et al. 2023). In people living with dementia (PLWD) sleep disturbances have been assessed by self-, carer-report and using wearables. In Alzheimer’s disease, the most common form of dementia, self-, carer-reported and actigraphically assessed sleep disturbances include early sleep timing, long time in bed, and nocturnal wandering to frequent interruptions of the nocturnal sleep period, long day time naps and almost complete absence of circadian organisation (Benca et al. 2022; Casagrande et al. 2022; Winsky-Sommerer et al. 2019). Understanding factors that drive the between and within PLWD variation in sleep disturbance may provide insights into mechanisms underlying associations between sleep disturbance, symptom severity and disease progression. Mechanistic insights may further help identify novel targets for improving sleep and reducing circadian disturbance in Alzheimer’s (Wang and Holtzman 2020; Balouch et al. 2022). Here we make use of a unique longitudinal cohort study in people living with dementia in the community. The longitudinal approach enables the detection of within participant variation, improving on approaches taken previously in the general population. Using interpretable digital health technology measurements along with information on outdoor and indoor environmental variables enables a comprehensive description of how nocturnal behaviour and physiological variables vary across seasons and associate with putative drivers of those changes.

Sleep is in part regulated by circadian rhythmicity which has evolved as an adaptation to the daily and seasonal variation in environmental variables including light and temperature (Bodenstein et al. 2012). In mammals, circadian rhythmicity in physiological variables such as body temperature, is driven by outputs of the circadian pacemaker located in the suprachiasmatic nuclei, the so-called ‘master-clock’ in the brain (Dijk and Czeisler 1995). This pacemaker is synchronised to the photoreceptive system in the eye. This photoreceptive system includes rods and cones and intrinsically photosensitive melanopsin expressing ganglion cells (Lucas et al. 2014) and there are direct connections from this photoreceptive system to the pacemaker. Light also has direct positive effects aspects on brain function including mood, alertness and cognitive performance (Gaggioni et al. 2014; LeGates, Fernandez, and Hattar 2014). Absence of light input, abnormally timed or reduced light exposure all lead to disturbance of sleep timing and waking function (Meyer et al. 2022).

Light exposure in humans is determined by the natural environmental light-dark cycle, human-made light and behavioural factors. In locations away from the equator, the external environmental light-dark cycle varies with season, and this drives seasonal changes in physiology and behaviour in many organisms (Lincoln 2019). Some of the seasonal changes in behaviours such as sleep, may also be driven by seasonal variation in environmental temperature. Circadian rhythmicity, sleep and temperature regulation are intimately related (Harding, Franks, and Wisden 2020). The 24 h variation in core body temperature parallels the environmental temperature rhythm with lowest values occurring close to wake time and sunrise respectively. Sleep lowers core body but extreme variations in environmental temperature pose a challenge to the regulation of body and brain temperature. High environmental temperatures, which due to climate change are becoming more common, disrupt sleep especially in vulnerable populations (Chevance et al. 2024).

In humans, compared to many other organisms, the magnitude of the seasonal variation in physiology and behaviour appears to be small. This may be related to the availability of human-made light and because modern humans spend most of their time indoors, the environment of which is to a larger extent invariant across seasons (Roenneberg 2004). Nevertheless, several recent cross-sectional studies have documented that in humans self-reported sleep duration is longer in winter than in summer (Zolfaghari et al. 2023; Titova et al. 2022; Suzuki et al. 2019). Studies on seasonal variation in sleep assessed objectively by actigraphy or PSG, are rare and often of a small scale. One notable exception is a cross-sectional study conducted in Japan in which objective sleep parameters were assessed in 68,604 participants and combined with meteorological data (Li et al. 2021). Not only did this study report seasonal variation in sleep duration, with a minimum during the summer, but also identified contributions of outdoor temperature to sleep quality.

There have been few longitudinal studies that consider the associations between the <u>indoor</u> light and temperature environment and sleep behaviour and none that capture day-to-day variation over long periods of time (Biller et al. 2024). Novel technologies such as contactless monitoring of bed occupancy, heart and breathing rate now enable longitudinal recording of sleep behaviours and physiology without burden to participants (Zhai et al. 2023; Boiko, Martinez Madrid, and Seepold 2023; Rigny et al. 2024). We analysed data obtained from an under the mattress digital health technology device to assess nocturnal behaviour, heart rate and breathing rate. The use of data from indoor light and temperature sensors enabled a detailed analysis of how environmental variables and physiological variables associate across seasons in people living with dementia who, just as other vulnerable groups, may be more sensitive to variations in the indoor environment.

Thus, the overall aim of this study was to assess seasonal variation in sleep behaviour, aspects of physiology, and a marker of the timing of the biological clock in people living with dementia in the community, and how these variations are modulated by light and temperature in their home environments. This focus on community-dwelling individuals is crucial as it represents the majority of people living with dementia, offers insights into more diverse environmental conditions than standardized care facilities, and provides an opportunity to inform practical, home-based interventions. By employing zero burden non-invasive monitoring technologies over an extended period, this study uniquely captures the complex interplay between environmental factors and sleep patterns in this vulnerable yet understudied population, potentially leading to improved sleep management and quality of life for people living with dementia in community settings.

## Results

### Selected participants and data

Seventy community dwelling participants (26 female, 44 male) from the MINDER clinical trial (IRAS number: 257561), which is a study designed to utilise health technologies in the home to support, monitor and manage the health of PLWD, were included in the analyses. Of the 70 participants 48 (69%) were diagnosed with Alzheimer’s disease, 19 (27%) with mild cognitive impairment or other dementias such as Parkinson’s disease or Lewy body dementia and a few (2, 3%) with other health conditions (frailty, stroke). For one participant the diagnosis was missing.

At baseline, participants were on average 79 years old (range 59-97) with mini-mental state examination (MMSE) scores that ranged from 2 to 30. Approximately half (36, 51%) had a MMSE score lower than or equal to 23, indicative of clinically significant cognitive impairment. A few participants (4, 6%) were severely impaired with MMSE scores lower than 10. Three participants were missing an MMSE assessment. Participant demographics are summarised in Supplementary Table S1.

The participants were located in the South of the UK, either in West London or in Surrey, living in their own homes which were equipped with multiple sensors. Light and temperature data in multiple rooms and bed occupancy, sleep efficiency, heart rate and breathing rate derived from an under the mattress ballistographic bedmat, which we validated in relevant populations (Ravindran et al. 2023a; Ravindran et al. 2023b; Ravindran et al. 2024) were collected between the 28^th^ May 2021 and the 19^th^ of June 2023. The total number of 24-h periods for which simultaneous measurements were available was 26,523. In each of the 70 participants there were at least seven days (24-h periods) of data available simultaneously from the lounge, bedroom and bedmat sensors in each of five of six possible two-month periods (Jan-Feb, Mar-Apr, May-Jun, Jul-Aug, Sep-Oct, Nov-Dec). Data were available on average 379 days / participant. For further details see Supplementary Table S1.

Indoor light and temperature data, bed occupancy, heart rate and breathing rate for two participants are shown in Fig. 1 along with light and temperature data from the external environment as reported by the UK Met office (Analysis 2019). Data are plotted in local clock time.

**Figure 1:**
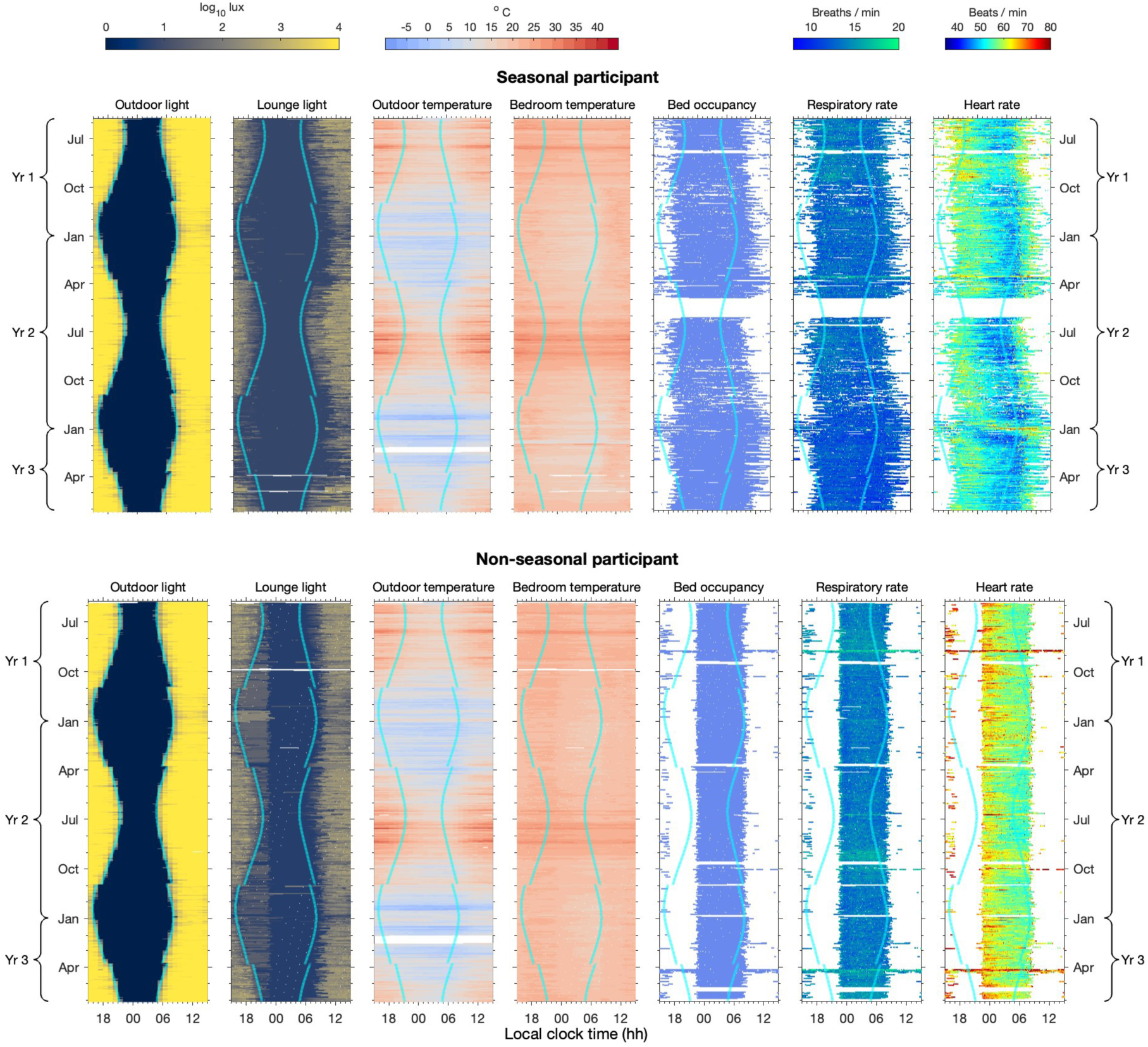
Seasonal variation in environmental and physiological variables for two participants. Annual patterns of outdoor and indoor light intensity, temperature, bed occupancy, respiratory rate, and heart rate for a seasonal (top) and non-seasonal (bottom) participant. Data span three consecutive years (Yr-1 to Yr-3), each panel representing a different variable. Each row depicts a single day. Colour scales indicate intensity or rate. Cyan curves denote sunrise and sunset times. Data are plotted in local clock time; discontinuities in sunrise/sunset lines reflect transitions between Standard Time and Daylight Saving Time.

### Observational patterns of indoor and outdoor environmental variables

Not surprisingly, the patterns of outdoor light and darkness align closely with the times of sunset and sunrise (Fig. 1). The large variation in the natural daylength (16h 38 min to 7h 50 min) at this latitude (51-52° N) is evident as are the transitions between Standard Time and Daylight Saving Time. Similarly, the outdoor temperature follows expected seasonal patterns, with warmer days in the summer than the winter and temperature dropping overnight. Occasional periods of particularly high temperatures can be seen in the summer, some of which were formally classified as ‘heatwaves’ according to the UK Met office.

For the two participants’ homes shown in Fig. 1, data from the multi-sensors show that seasonal changes in the outdoor light and temperature environment are to some extent echoed by seasonal changes in the indoor light and temperature environment. For light, the seasonal pattern of changes in the natural photoperiod are overlaid by two further contributing factors. First, after sunset, the indoor light environment is brighter than the outdoor light environment likely due to electric light. Second, light intensity in the lounge remains low until participants get up in the morning, likely due the effect of blinds and / or curtains. For temperature, the daily pattern of cooler bedrooms at night than during the day and the seasonal pattern of cooler bedrooms during the winter than the summer reflect the daily and seasonal outdoor temperature changes. But the amplitude of both the daily and seasonal changes are reduced. It is particularly evident that during the hottest periods, bedrooms remain hot overnight even when the outdoor temperature drops.

### Factors influencing indoor light and temperature dynamics

Multiple factors dictate the internal light and temperature environment including the fact that we can exert some control over these variables. To identify dominant drivers of indoor light and temperature, hourly light and temperature values (∼3.15 million values for each of light and temperature) were analysed with a fixed effect model. The model incorporated hour of the day, season, room and home as fixed effects. “Season” was categorised by two-month periods. The “rooms” included were the lounge, bedroom, bathroom and kitchen. All 70 participant homes were included. All effects were significant (p < 0.0001 in all cases), but while hour and season had the largest effect sizes for light, home and season had the largest effect size for temperature, see Supplementary Table S2. Daily and seasonal differences in light and temperature are illustrated for example homes in Fig. 2 (a).

**Figure 2:**
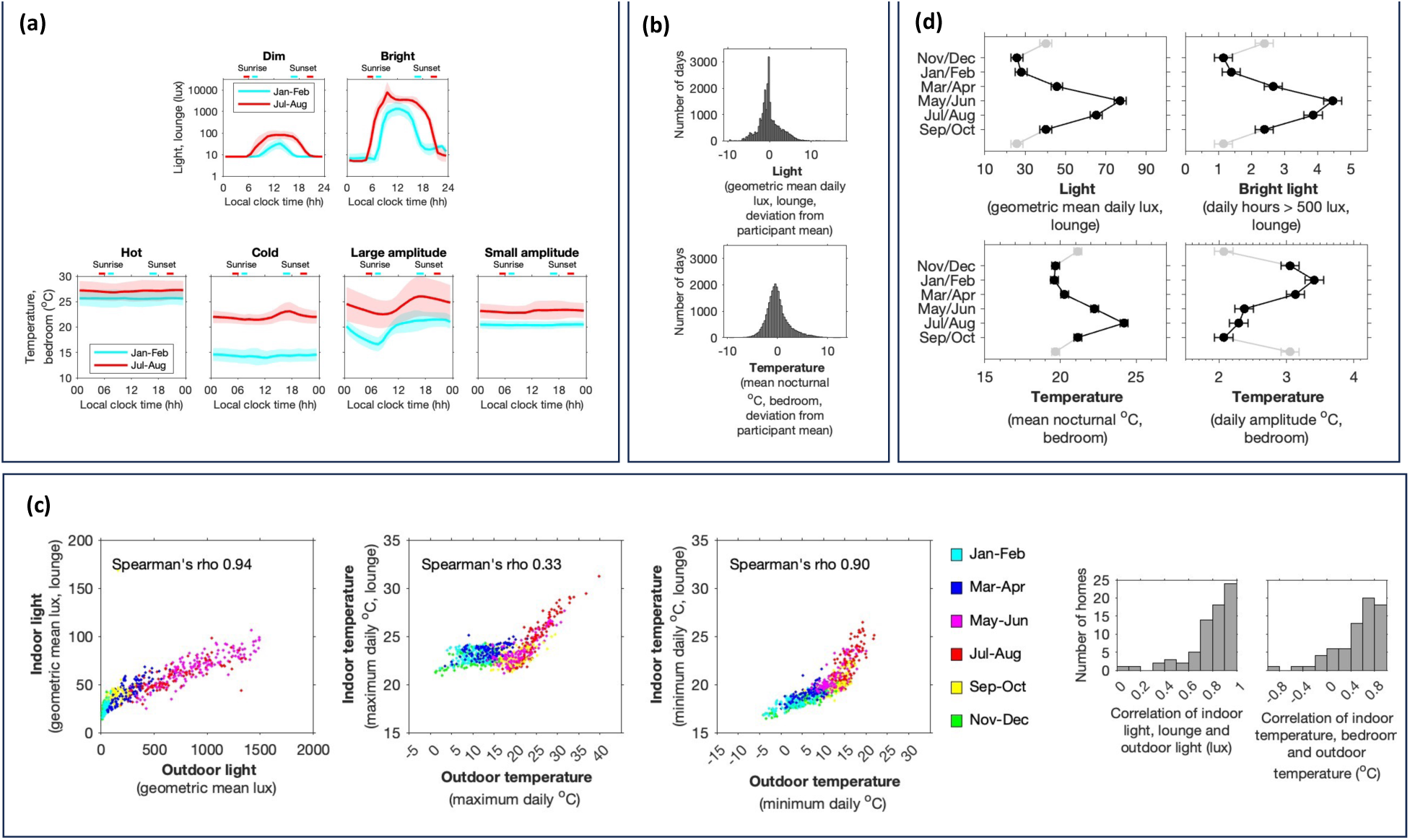
Daily and seasonal variation in the indoor light and temperature variation and association with the outdoor light and temperature environment. (a) Daily and seasonal variation in light (two homes) and temperature (four homes). Homes were selected to illustrate the observed range of light and temperature environments. All data were plotted in local clock time. (b) Deviation from participant mean for average light levels in the lounge and nocturnal bedroom temperatures. (c) Associations between the indoor and outdoor daily light and temperature environment. Each data point represents one calendar day. For each calendar day, an indoor average daily value was found by averaging across all homes. The outdoor average daily value was found from the MET office data. Also shown are histograms showing the strength of the indoor to outdoor association for each participant. (d) Least squares (LS) means for the seasonal variation of daily measures of the indoor light and temperature environment calculated from a linear mixed effect model with home as a random effect.

An analysis of the within and between home variation in average daily indoor temperature and light revealed that the within home variance was slightly higher than the between home variation with intraclass correlation coefficients (ICCs) of 0.44 for hours of bright light in the lounge and 0.47 for mean nocturnal temperature in the bedroom Supplementary Fig. S2. The distributions of the within home (participant) variation in the average light levels in the lounge and nocturnal bedroom temperatures are plotted in Fig. 2 (b).

Across all homes, average indoor light and temperature were strongly associated with outdoor temperature and light, see Fig. 2 (c), even though average indoor light levels were substantially lower than average outdoor light levels. These values suggest that the geometric mean indoor light levels were typically 20 times lower than the geometric mean outdoor light levels. For temperature, the association between the outdoor and indoor values is to a larger extent driven by high temperatures: when the maximum outdoor temperature fell below approximately 20 C°, indoor temperatures were typically maintained at between 20 C° and 25 °C. The strength of the correlation between the indoor and outdoor environment varied by home, with indoor light more consistently correlated with outdoor light than temperature, see Fig. 2 (c).

To relate environmental factors to *daily* sleep behaviour (e.g. duration of time in bed, sleep efficiency), three *daily* measures of the light and temperature environment were constructed (hours of light over 500 lux, mean nocturnal temperature, daily temperature amplitude). We elected to describe temperature using mean and amplitude because minimum and maximum temperature, are highly (Spearman’s rho 0.84), whereas mean and amplitude are only weakly correlated (Spearman’s rho –0.24). As illustrated for the “high amplitude” home in Fig. 2(a), high amplitude largely reflects the overnight drop in temperature. Linear mixed effect models with home as a random effect showed that there were significant effects of season on all of the daily light and temperature measures (p<0.0001 in all cases) (see Supplementary Table S3, and Fig. 2 (d). Consistent with seasonal variation in the outdoor environment, recorded light levels and temperatures were higher in the summer months. The amplitude of the change in temperature (daily maximum – daily minimum) is greatest in the winter.

### Average duration, timing and fragmentation of in-bed occupancy, breathing and heart rate

In Fig. 1, the data obtained from the bedmat sensor highlight some of the large differences in bed occupancy patterns between and within individuals. In participant ‘seasonal’ the bed was occupied for much of the time (on average 12:43 hh:mm +/-3:35 h:mm per 24 h) but was disrupted with frequent periods during the night when the participant left the bed. Although there was a period of approximately a month in May at the end of the first year when no data were recorded, bed occupancy appears to be seasonal with longer time in bed during the winter months. In contrast, participant ‘non-seasonal’ had a very regular and shorter (on average 9:07 h:mm +/-0:58 h:mm per 24 h) pattern of bed occupancy.

For both participants shown in Fig. 1, the physiological recordings show that the heart rate drops to a minimum value in the early hours of the morning, rising again prior to the morning bed exit time. In the case of the ‘seasonal’ participant, there appears to be a period at the end of year 2 / beginning of year 3 when the time of the minimum of heart rate first drifts later and then appears to desynchronise from the 24-h day. For a further analysis of this desynchrony effect, see the Supplementary material.

The fragmented nature of the in-bed time (and corresponding sleep epochs as detected by the bedmat algorithm) for some participants implies that it is not straightforward to define commonly used metrics of sleep behaviour such as sleep onset, sleep offset and duration. We therefore took the approach of defining three duration measures of in-bed occupancy (total number of minutes for each 24 h period in which the bed was occupied, duration of the major nocturnal in-bed period, duration of any in-bed periods initiated during the day) along with associated timing measures. In addition, we considered measures of fragmentation (e.g. number of bed exits / hour, wake after sleep onset, sleep efficiency) and measures of physiology (e.g. heart rate and breathing rate). Since the timing of the minimum heart rate can provide information on the diurnal variation in autonomic function and has been used as a marker of circadian phase (Vandewalle et al. 2007; Bowman et al. 2021; Natarajan et al. 2025) this was also one of the measures we evaluated. In total, we considered 25 different measures of duration, timing and fragmentation of in-bed occupancy, physiology and a marker of the timing of the biological clock. Further detail is given in the methods. Definitions of all 25 measures are given in material section S2. The correlation between the 25 different measures is shown in Table 1. In general, there were moderate to strong associations between duration and timing measures for the major nocturnal in-bed periods. In addition, many of the measures of fragmentation showed a significant correlation with each other, for example, bed exit rate was associated with the median duration of sleep bouts (Spearman’s rho –0.38). In our study, we elected to report on all measures to present a detailed picture as to time-in-bed behaviour and highlight how commonly reported sleep metrics are related to each other.

**Table 1:**
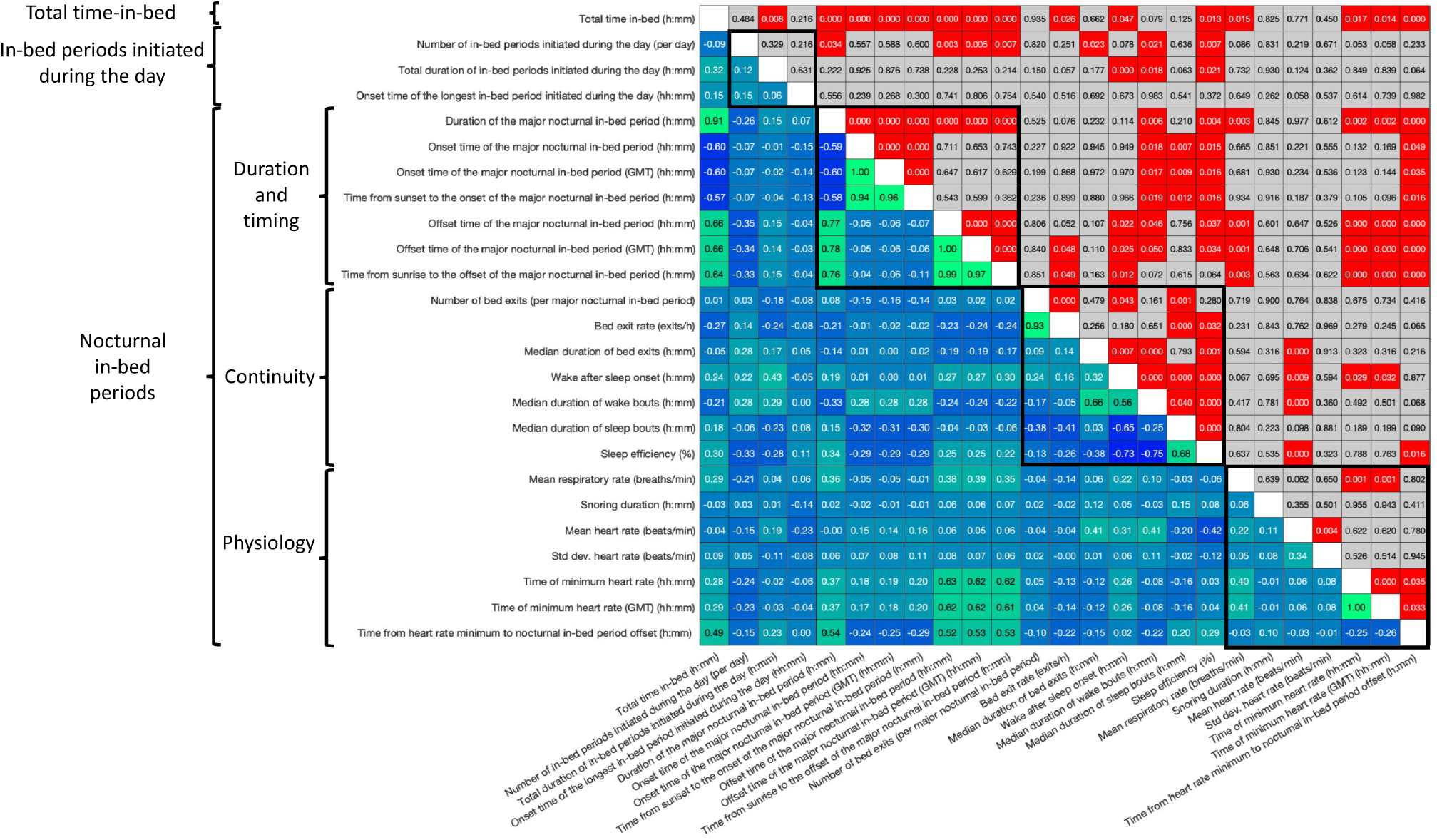
Correlations between duration, timing and fragmentation of bed occupancy and sleep physiology measures. Correlation coefficients (Spearman’s rho) and p values for the 25 measures of sleep behaviour and physiology. The correlations were calculated as the correlation between participant mean values (n=70).

On average, participants spent 8:48 h:mm +/-2:05 h:mm in bed in each 24 h period. The average duration of major nocturnal in-bed periods was 8:47 h:mm +/-2:09 h:mm, starting at 23:05 hh:mm +/-1:18 h:mm and ending at 07:51 h:mm +/-1:33 h:mm (see Supplementary Table S4). Participants typically went to bed 4:28 h:mm +/-1:19 h:mm after sunset and got out of bed 1:19 h:mm +/-1:35 h:mm after sunrise. The values reported here are means +/-standard deviation for further details and measures (e.g. standard error, median, minimum, maximum) see Supplementary Table S4.

During each major nocturnal in-bed period, participants on average spent 1:15 h:mm +/-0:41 h:mm awake after sleep onset and got out of bed 0.4 +/-0.37 times / h. Most periods out of bed were short (median duration 0:06 h:mm +/-0:04 h:mm per nocturnal in-bed period). For the majority of participants (46 out of 70, 66%) the major nocturnal in-bed period was on average longer on the weekend nights of Friday and Saturday (0:11 h:mm +/-0:27 h:mm), starting 0:06 +/-15 h:mm +/-later and ending 0:16 h:mm +/-26 h:mm later, see Supplementary Fig. S3.

Participants spent 0:17 h:mm +/-0:21 h:mm snoring and had an average heart rate of 59.47 +/-6.69 beats/min and a mean respiratory rate of 14.89 +/-2.16 breaths/min (see Supplementary Table S4).

The major nocturnal in-bed period was supplemented by minor nocturnal in-bed episodes and, occasionally, in-bed periods that were initiated during the day (mean 0.14 +/-0.24 per day). When in-bed periods were initiated during the day (i.e. naps), the mean duration was 1:39 h:mm +/-0:57 h:mm (see Supplementary Table S4).

Fig. 1 highlights the data for two participants. In general, there were large variations within participants, indeed 20 out of 25 of the measures we considered had intraclass correlation coefficients less than 0.5 (see Supplementary Table S4) indicating that the within participant variation was larger than the between participant variation. Only mean heart rate and respiratory rate had ICC’s which were substantially greater than 0.5 (0.80 and 0.85 respectively), indicating that heart and breathing rate are more trait-like. Boxplots contrasting between and within variation in measures representing time-in-bed and physiology are shown in Supplementary Fig. S2.

Differences between participants in most measures showed only weak associations with MMSE, NPI and age. There was a weak to moderate association (Spearman’s rho 0.35) of the time of the onset of the major nocturnal in-bed period, such that those with a lower MMSE went to bed earlier, see Supplementary Table S5. Participants with hotter bedrooms at night had more fragmented sleep and had more in-bed periods initiated during the day (naps).

### Seasonal changes in duration, timing and fragmentation of in-bed occupancy, physiology and a marker of the biological clock

We assessed seasonal effects on the 25 measures of in-bed occupancy, physiology and markers of the biological clock using linear mixed effect models in which we controlled for gender, age on entrance to the study and diagnosis (Alzheimer’s vs not Alzheimer’s) with participant as the random effect. We observed significant effects of season on 22 out of the 25 variables analysed. Results are summarised in Supplementary Table S6, Supplementary Fig. S4 and in Fig. 3 (a).

**Figure 3:**
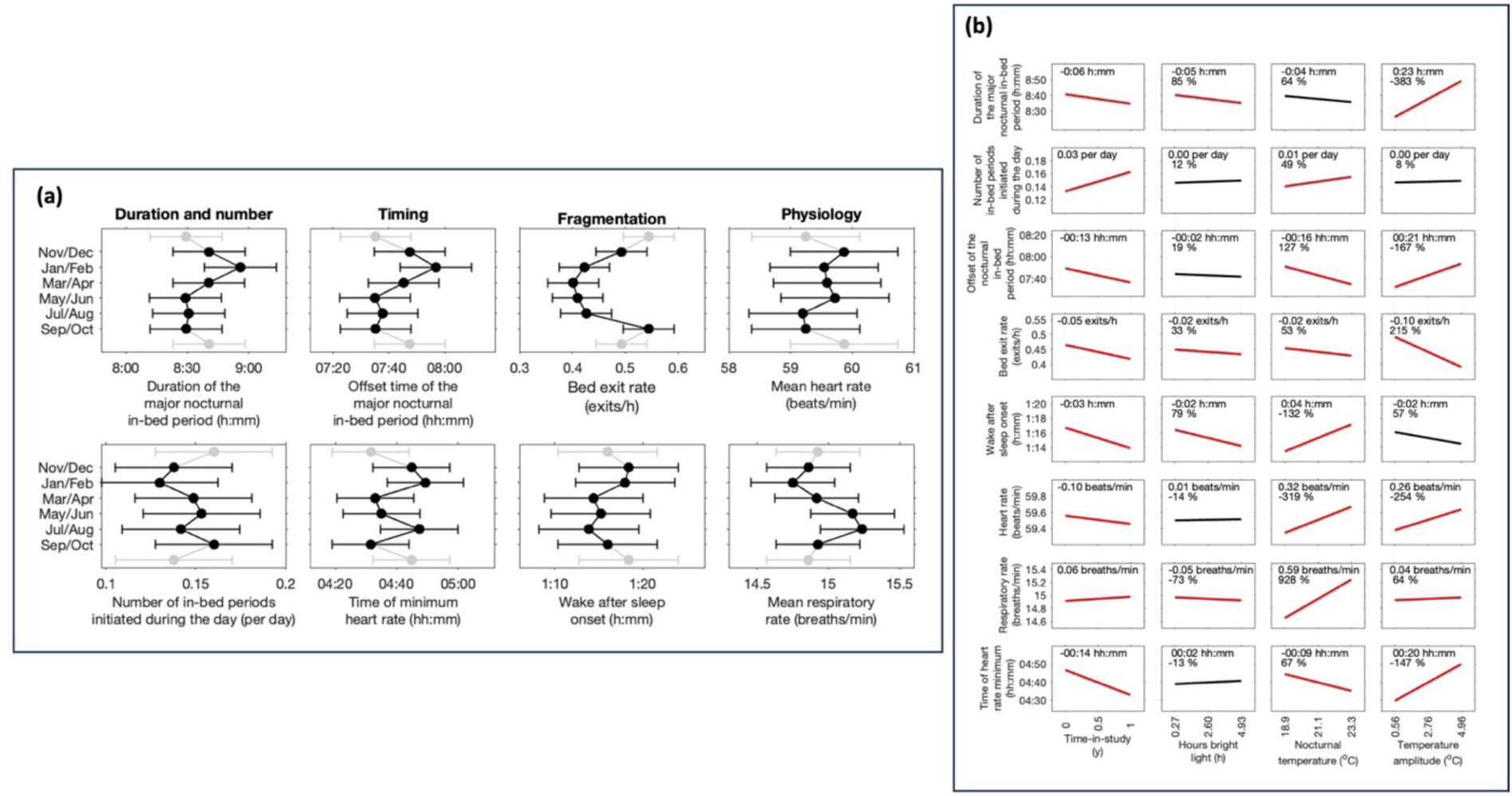
Seasonal effects on the duration, timing and fragmentation of bed occupancy and on physiology. (a) Seasonal variation in 8 measures of duration, timing, fragmentation and physiology. The data shown are the Least Squares (LS) means from a linear mixed effect model which considers the seasonal variation for each measure controlling for gender, age and diagnosis with participant as a random effect (see Supplementary Table S6). Equivalent LS means graphs for all other measures considered are shown in Supplementary Fig. S4. (b) The predicted effects of one year in the study and variation in light and temperature over the mean +/-one standard deviation for a typical participant. The predictions come from the effect sizes in the linear mixed effect models which consider seasonal variation in in-bed behaviour and physiology along with environmental light and temperature controlling for gender, age, diagnosis and time-in-study with participant as a random effect as reported in Supplementary Table S7. The significant effects are shown in red. In each panel, the numerical values indicate the change over the range that is shown. The percentages indicate the percentage change relative to the predicted effect of change of one year in the study. For example, the effect of temperature on nocturnal time-in-bed is 3.83 times the effect of one year in the study in magnitude and in the opposite direction.

### Seasonal effects on duration and timing of in-bed occupancy

The duration of the major nocturnal sleep period varied significantly with season from 8:55 h:mm in Jan-Feb to 8:28 h:mm in the summer months, see Fig. 3 (a), primarily as a consequence of getting up later in winter. Even with the large changes in natural photoperiod through the year, people usually went to bed after sunset and got up after sunrise. There was no significant seasonal effect on the start of the major nocturnal time in-bed period in local clock time. Analysing onset and offset time in GMT rather than local clock time, increased the magnitude of the seasonal change and introduced significant seasonal differences for both onset and offset times (see Supplementary Fig. S4). When in-bed periods during the day occurred, there was no significant seasonal difference in the duration of the in-bed periods, but there were significantly more in-bed periods in the summer than in the winter.

There was no significant difference between those with and without an Alzheimer diagnosis in duration or in-bed timing measures relative to the 24 h day. Interestingly, the time of the heart rate minimum occurred significantly closer to the time at which people got up in those without an Alzheimer’s diagnosis.

### Seasonal effects on in-bed fragmentation

Seven measures of in-bed fragmentation during the major nocturnal in-bed period were considered: number of bed exits, bed exit rate, median duration of bed exits, wake after sleep onset, median duration of sleep bouts, median duration of wake bouts and sleep efficiency. All seven measures varied significantly with season indicating that sleep was less interrupted in the spring and summer than autumn and winter. For example, bed exit rate was lowest in the spring (Mar-Apr) and highest in the autumn (Sep-Oct). When people got up more often, the duration of the out of bed bouts was on average shorter. Men got out of bed less often than women and had a lower bed exit rate.

### Seasonal effects on physiology and the timing of the minimum of heart rate, an indicator of the biological clock

Both heart rate and respiratory rate varied significantly with season. Heart rate was fastest in the winter months of Nov/Dec and slowest in the summer months of Jul/Aug. Whereas respiratory rate was slower during the winter than the summer. Those who were older had a significantly faster heart rate and shorter snoring duration. The timing of the minimum heart rate varied significantly across season with a bimodal pattern with later timing in both Jan/Feb and July August (see Fig. 3). When analysed in GMT rather than local clock time the bimodality in the timing of the minimum heart rate disappeared (see Supplementary Fig. S4). The time from the heart rate minimum to the end of the nocturnal in-bed period was shorter in the summer than in the winter.

### Light and temperature affect duration, timing and fragmentation of in-bed occupancy, and physiology even when controlling for seasonal effects

To assess whether variation in the indoor light and temperature environment had an effect on the same 25 measures of in-bed occupancy, physiology and markers of the biological clock we extended the linear mixed effect model to include fixed effects of hours of bright light (light > 500 lux) in the lounge, mean nocturnal bedroom temperature and the temperature amplitude in the bedroom controlling for season (two-month period), gender, age on entrance to the study, time in study and diagnosis (Alzheimer’s vs non Alzheimer’s). Participant was considered as a random effect. Results for time in study, light and temperature are summarised in Supplementary Table S7 and in Fig. 3 (b).

### Effect of Time in Study

In dementia, changes in sleep over time may be an important indicator of deterioration in cognitive function and / or ability for carers to manage at home. Nearly all measures (22 out of 25) showed a significant time-in-study effect, with longer time-in-study associated with more in-bed periods initiated during the day, longer in-bed periods initiated during the day but shorter duration of the major nocturnal in-bed period. With longer time-in-study, the major nocturnal in-bed period started and finished earlier, and the time of the heart rate minimum was earlier. Bed exit rate went down and wake after sleep onset became shorter, but at the same time there were indications of more disrupted sleep with the median duration of wake bouts going up and sleep efficiency going down with time in study.

### Effect of light

Over and above the effects of seasonal, more hours of bright indoor light were associated with significantly shorter total time in-bed (0:05 mins shorter, 85% of the predicted one year change), shorter major nocturnal in-bed periods, and decreased fragmentation of the nocturnal sleep period as indexed by fewer bed exits, a reduced bed exit rate (rate reduced by 0.02 exits/h, 33% of the predicted one year change), shorter duration of bed exits, and less wake after sleep onset. More hours of bright indoor light were also associated with lower breathing rate, reversing the annual trend (predicted one year change is an increase in 0.06 breaths / min, the effect of light is a predicted decrease by 0.05 breaths/min). In addition, more light was associated with longer snoring duration, and reduced standard deviation in heart rate. Interestingly, we found that the number of hours of bright light did not significantly alter the timing of the heart rate minimum, although more hours of bright light were associated with a shorter interval between the heart rate minimum and the end of the major nocturnal in-bed period.

### Effect of temperature

Over and above seasonal effects, nights with hotter bedroom temperatures were significantly associated with people going to bed earlier, getting up earlier (16 mins earlier, 127% of the predicted one-year change) with an earlier time of the heart rate minimum (9 mins earlier, 67% of the predicted one year change), see Supplementary Table S7. On hotter nights, people got out of bed significantly fewer times, but when they did get up, they got up for longer, and wake bouts and wake after sleep onset were longer. Overall sleep efficiency was lower when nights were hotter. Mean heart rate and respiratory rate were significantly raised (–319% and –928% of the predicted one-year change respectively), although snoring duration went down.

A higher amplitude of the daily indoor variation in temperature, which primarily reflects lower bedroom temperatures at night, significantly increased time in-bed duration measures, increasing the duration of the nocturnal in-bed period by 23 mins (–383% of the predicted one-year change), primarily because rise times were later (later by 21 mins, –167% of the predicted one year change). The time of the heart rate minimum also shifted later by a similar amount (later by 20 mins, –147% of the predicted one-year change). The number of bed exits and bed exit rate was lower (reduced by 0.1 exit/h, 215% of the predicted one year change) and sleep efficiency increased with higher amplitude of the daily variation in temperature. Mean heart rate and respiratory rate were again significantly raised (–254% and –64% of the predicted one-year change respectively), as was the standard deviation in heart rate.

### Differences in seasonality between participants

There were large differences between individuals in the magnitude of the seasonal effect. Two examples are shown in see Fig. 4(a), with all remaining participants shown in Supplementary Fig. S5. To quantify the degree of seasonality, for each participant we considered the difference between the mean duration, onset and offset of the major nocturnal in-bed period in the winter (Jan-Feb) and the summer (Jul-Aug). The “seasonal” participant shown in Fig. 4 (a) ranks in the top quartile for the magnitude of the seasonal changes, while the “non-seasonal” participant ranks in the third quartile for the magnitude of the seasonal changes. Histograms showing the size of the seasonal effect for the duration, onset and offset of the major nocturnal in-bed period are shown in Fig. 4(b). The seasonality of participants was not significantly associated with age, MMSE or NPI or seasonal changes in temperature. However, there was a significant association with seasonal changes in indoor light with seasonal differences in indoor light explaining approximately 12% of the variance. There was also a significant association between weekend – weekday differences in the time people got out of bed and seasonal changes in onset time, see Fig. 4(c) and Supplementary Table S8.

**Figure 4:**
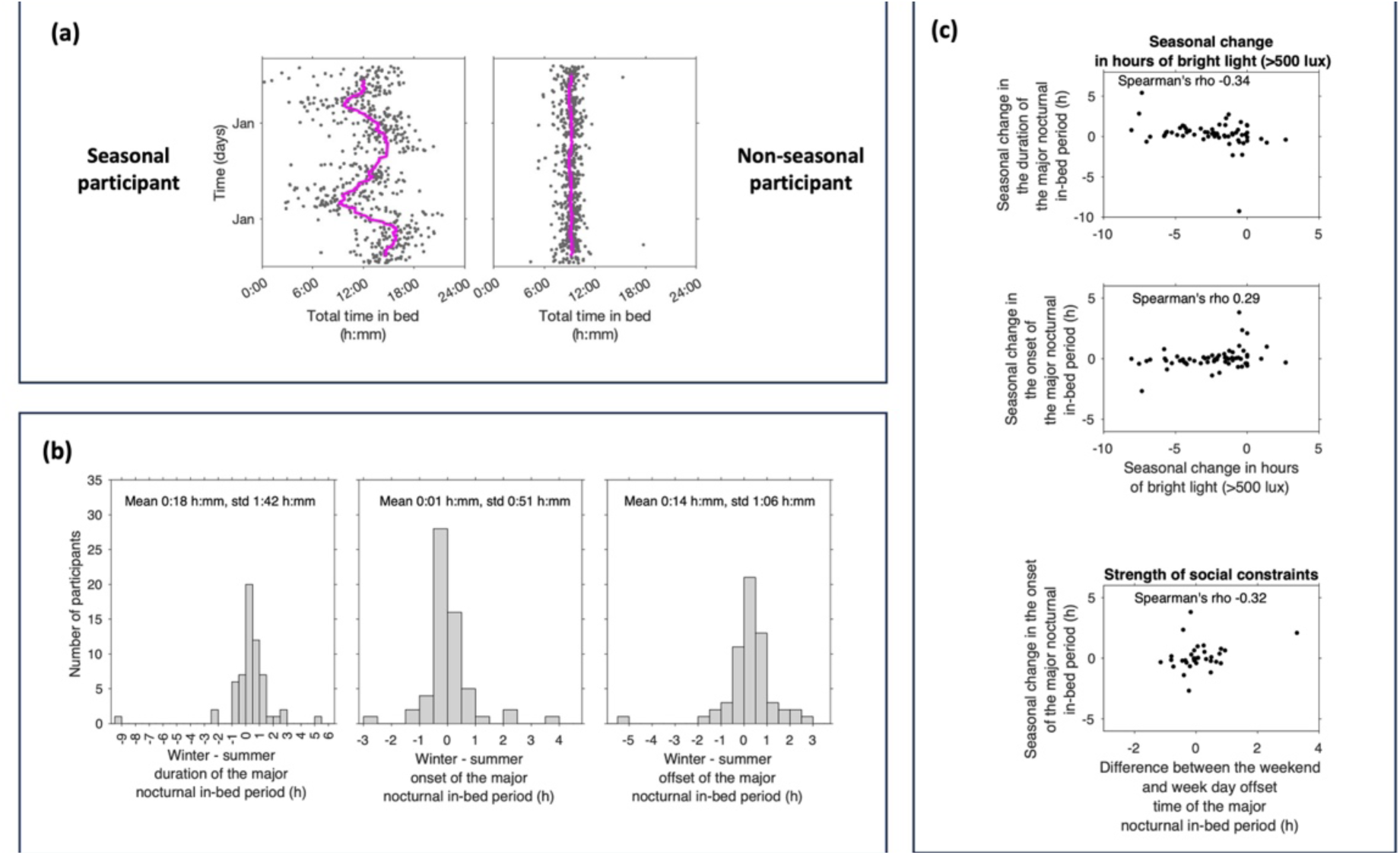
Quantifying seasonality and drivers of seasonality between participants. (a) Illustrative examples showing the time course of the total time in-bed for the two participants used in Fig. 1. Each data point is the value for one day. The magenta lines show the two-month moving averages. (b) Histograms of participant seasonality in duration, onset and offset of the major nocturnal in-bed period per participant (n=70). Seasonality was calculated as the difference between the participant mean in the winter (Jan-Feb) minus the participant mean in the summer (Jul-Aug). (c) Correlations between selected possible correlates of seasonal change and the duration and time of onset of the major nocturnal in-bed period.

## Discussion

Longitudinal objective assessment of sleep behaviour and physiology with zero-burden digital health technology demonstrated that sleep timing and fragmentation as well as heart rate and breathing rate all vary with season in people with dementia who are living independently at home. Simultaneous assessment of the indoor light and temperature environment revealed that these variables also vary with season and that beyond the effect of season, lower indoor light during the daytime and higher temperature during the nighttime associate with more disturbed sleep. Indoor light and temperature are modifiable factors that can be targeted to reduce sleep disturbance in people living with dementia.

Indeed, recent recommendations are that daytime indoor light levels should be at least 250 melanopic lux(Brown et al. 2022) which for typical room lighting means more than 500 lux (Schlangen and Price 2021). Modelling of the human sleep-circadian timing system suggest that this may be conservative estimate of the levels needed for appropriate and robust synchronisation of the circadian clock with the environment, i.e. entrainment (Skeldon, Phillips, and Dijk 2017). Typical light levels in homes were measured to be well below these recommended levels, leaving considerable scope for improvement. For example, supplementing lighting during the day and reducing lighting levels during the evening can significantly improve synchronisation of the circadian clock and sleep and health outcomes. Large epidemiological studies have indeed shown that appropriate light-dark cycles are associated with mood (Burns et al. 2021), mental health (Yu et al. 2022) and reduced mortality risk (Windred et al. 2024).

Commonly reported recommendations for bedroom temperature are in the range 15°C to 20°C, although the scientific basis for these levels is unclear. The current UK bedroom threshold delineating overheated bedrooms is 26°C (Lomas and Li 2023). A recent study in an older population found a u-shaped relationship between overnight temperatures and sleep efficiency, with reduced efficiency when temperatures were too low or too high. Overnight temperatures in the range 20°C to 25°C resulted in highest sleep efficiency, with optimal temperatures dependent on the individual (Baniassadi et al. 2023). The mean overnight temperature observed in our study of 21.1°C is in line with these levels with approximately 10% of nights above 25°C. Sleep efficiency increased at lower temperatures and when the diurnal temperature amplitude was higher. Advice has frequently focussed on methods to keep bedrooms cool during periods of hot weather such as having windows open overnight, keeping curtains / blinds shut during the day when bedrooms are unoccupied. We found that some homes were uniformly hot throughout the day and night and at all times of year, suggesting there is scope for more general advice on turning down thermostats overnight also in people living with dementia.

The observed seasonal variation in the nocturnal in-bed period with duration 27 mins longer later and rise times delayed by 22 minutes in the winter in PLWD is in accordance and comparable in magnitude with previous results obtained from self-report and actigraphy data in the general population for sleep duration. For example, a longitudinal study reported 16.8 min longer sleep in the winter for a population with depression (Zhang et al. 2024); and in a large cross-sectional study of a Japanese cohort (age 10-89 y) accelerometer derived sleep time (but not in-bed time) was 39.6 min longer in the winter, primarily driven by later rise times (Li et al. 2021). This suggests that in PLWD ‘season’ affects measures of duration and timing of sleep as it does in the general population.

The longitudinal and objective sleep assessment approach of the current study and identified within individual seasonal responsiveness is unique certainly for PLWD. Our results highlighted that sleep fragmentation, which was often not assessed in previous studies, is particularly sensitive to seasonal, light and temperature effects. This is relevant because sleep fragmentation is associated with the risk of dementia (Lysen et al. 2020). Furthermore, night-to-night variation in sleep fragmentation associates with day-to-day variation every day memory errors, behavioural problems and alertness in people living with Alzheimer’s (Balouch et al. 2022).

Interestingly, two recent large scale cross-sectional studies (Mooldijk et al. 2022) (n=10,276) and (Lim et al. 2018) (n=3,353 participants) have reported seasonal variation in cognition and their relevance to dementia. In both studies cognition varied significantly with season with peak performance in the summer. Furthermore, dementia diagnoses and the odds of meeting criteria for mild cognitive or dementia also varied with season with a maximum in winter/spring. Factors driving this seasonal variation in cognition and dementia diagnoses, have not been identified although putative drivers are seasonal variation in exposure to antigens, light, and seasonal variation in the sleep-wake cycle. Similarly, seasonal variation has also been reported in psychiatric symptoms and diagnoses such as seasonal affective disorder (Zhang and Volkow 2023) in humans living in industrialised societies.

Our approach highlighted that in the summer when sleep is short there were more in-bed periods initiated during the day. This is consistent with laboratory protocols in which it has been shown that shorter in bed periods lead to more consolidated sleep, but sleep that is too short results in an increase in daytime sleep propensity (Skorucak et al. 2018; Klerman and Dijk 2005). Such an increase in daytime sleep propensity in people living in the community may be expressed as more daytime naps. Together these observations underline the importance of the ‘right’ amount of sleep and are consistent with a recent pilot study indicating that restricting time-in-bed in healthy older adults can decrease sleep fragmentation (Wilckens et al. 2024).

The seasonal variation in heart rate and breathing rate during sleep is to our knowledge a novel observation. Mechanisms driving this variation can be manifold but may be related to seasonal variation in other environmental factors such as humidity and air quality. Alternatively, it may be related to seasonal variation in common mild ill health conditions (e.g. a cold) which may involve respiratory inflammation and congestion. Finally, it cannot be excluded that seasonal variation in heart and breathing rate is related to seasonal variation in the circadian modulation of these variables (Vandewalle et al. 2007; Hayter et al. 2021; Spengler, Czeisler, and Shea 2000; Butler et al. 2015). In this context it is of interest that the timing of the minimum of heart rate, which may be a phase marker of endogenous circadian rhythmicity varied with season and in a way that could not simply be explained by seasonal variation in sleep timing. This was most clearly demonstrated in the case study of desynchrony (see the Supplementary material section S5) but was also evident from the changed phase relationships between the timing of the minimum of heart rate and the end of the nocturnal time in bed period.

It is often implied that seasonal variation in physiology and behaviour in humans is driven by variation in the natural, i.e. outdoor environment which may appear to be at odds with the observation that most people in industrialized societies spend 90% of their time indoors (Klepeis et al. 2001). Indeed, previous population studies linking high temperatures to disrupted sleep have largely been based on weather station measurements of the external temperature environment (Obradovich et al. 2017; Chevance et al. 2024; Li et al. 2021). The present data demonstrate that the indoor light and temperature environment also vary seasonally. In fact, both indoor temperature and light correlated with outdoor temperature and light across and within season. Our findings for temperature mirror those found in a study carried out in the USA (Nguyen, Schwartz, and Dockery 2014) and emphasize the relevance of the indoor environment, and not only the outdoor environment, as a determinant of physiology and behaviour in general but also in PLWD. Indeed, our results demonstrate that even controlling for season, indoor light and particularly the indoor temperature environment had a significant effect on many aspects of in-bed occupancy and physiology. Between participants, we find that those who have habitually hotter bedrooms have more fragmented sleep. Within individuals, and consistent with previous studies e.g. (Li et al. 2021; Lan et al. 2017) we find that lower nocturnal bedroom temperatures were associated with lower sleep fragmentation. Within participants with cognitive impairments, previous work has shown that decreased objective measures of sleep fragmentation are associated with reduced memory errors and fewer behavioural problems (Balouch et al. 2022). Since we have considerable control over the indoor temperature environment, keeping bedrooms cool at night may be a pragmatic way to ameliorate some of the symptoms related to dementia. Designing homes and bedrooms to allow adequate temperature control may become even more relevant in the light of climate changes, as highlighted in a recent protocol paper (Lomas et al. 2024), particularly in geographical areas that are most affected.

The focus on the outdoor environment as the driver has led to active debate on the relative importance of seasonal changes in the timing of solar noon induced by Daylight Saving Time to those of societal constraints (e.g. (Skeldon and Dijk 2021)) particularly in early to mid-adulthood when work may impose constraints on sleep timing (e.g. (Shochat et al. 2019)). For this reason, all timing variables were analysed in both clock time and in standard time (GMT). In accordance with previous analyses (Skeldon and Dijk 2021), the present results suggest that the time at which people go to bed is dictated by clock time rather than the timing of sunset or the timing of the midpoint of the photoperiod. The latter time indicator varies by an hour when we shift between standard time (GMT) and British Summer Time (GMT+1) environmental factors. The only indication that the shift of GMT to BST has a lasting effect (i.e. longer than a few weeks) on the timing of physiological variables is the bimodal seasonal variation in the timing of the heart rate minimum.

## Limitations

In this study we investigated seasonal variation and variation in indoor light and temperature, but many other potentially important environmental factors such as humidity, air quality, time spent outside and weather were not assessed. Information on acute health events, e.g. a cold, which together with immune function may vary with season (Wyse et al. 2021), were not included in the analyses, in part because this information was not available. Information on seasonal and day-to-day variation in social factors, were also not considered although it is of interest that there were systematic changes between weekdays and weekends in timing of bed occupancy, but typically much smaller in magnitude than seen in younger populations (Roenneberg et al. 2012), and some people had “reverse” social jetlag in the sense that they went to bed earlier and / or got out of bed earlier at the weekend than during the week.

A strength of the study is the heterogeneity of the population and its longitudinal nature. A main focus of our linear mixed effect modelling was therefore the within participant changes. More granular data on changes with season on health, daily routines and obligations, use of alarm clocks and exposure to outdoor light could provide more understanding of what drives seasonal changes. We found some differences between men and women, but do not know how the overall health of man and women in our study compare.

The available data and approach could demonstrate associations between variation in environmental variables and sleep disturbance but could not show whether these associations extend to symptoms such as behavioural problems, reduced alertness, and everyday memory problems because in the protocol these data were not collected with a sufficiently high temporal resolution. The validity of the sensors used to collect information about the environment has been assessed to some extent (see supplemental materials), but the placement of these sensors varied across houses and may not always have been ideal. Furthermore, light was quantified in lux, which from a circadian-light perspective is not ideal. The spectral composition of light exposure varies with season (Thorne et al. 2009). Future studies should quantify either the spectral composition of light or at least quantify light as per current recommendations, i.e. as melanopic lux/Daylight Equivalent.

Although we have demonstrated that the measures calculated from the bedmat and reported in this study are valid in this population, the version of the bedmat used in the present study did not provide information on heart rate variability, did not quantify sleep apnoea and the sleep stage identification provided by this device is not accurate. Future studies should address these limitations.

## Conclusion and outlook

Integrating information about the indoor environment with information about sleep behaviour and physiology demonstrated the impact of variation in the indoor environment on sleep disturbance in dementia. This integration, enabled by longitudinal recordings through digital health technology, represents a powerful approach to identify both effects at the group level but also individual differences in the response to the environment. Understanding the contribution of indoor environmental variables to variations in behaviour and physiology may lead to interventions to improve sleep and reduce circadian disturbance in PLWD and enable them to live independently for longer. Since sleep disturbances are associated with negative health outcomes not only in dementia these approaches also hold promise for improving health in other segments of the population.

## Methods

### Protocol, ethics and participants

The data analysed for this report were collected in the MINDER Health Management Study for Dementia (IRAS number: 257561). MINDER seeks to support the management of the health and well-being of PLWD who are living in the community through the use of multiple sensors, self-reported measures, clinical evaluations and integration of this information. One of the objectives of MINDER is to evaluate and refine a suite of technologies to produce a scalable health management system. MINDER is a follow up and extension of the pioneering TIHM (Technology Integrated Health Management) study (ISRCTN46184518).

Participants have been continuously enrolled to MINDER since 2019 and is still ongoing. Most participants have received a diagnosis of dementia, including Alzheimer’s disease, mild cognitive impairment and vascular dementia. Some participants have a diagnosis of general frailty or other neurodegenerative conditions. Inclusion criteria included 50 years of age or older and at baseline sufficient functional English to complete assessment instruments. Exclusion criteria included treatment for terminal illness at baseline, severe mental health issues and suicidal thoughts. Participants in MINDER were assessed according to Good Clinical Practice guidelines and the Mental Capacity Act 2005. If they were able, they provided written informed consent otherwise a study partner acted as their consultee representative. The study was approved by the Health Research Authority’s London-Surrey Borders Research Ethics Committee (19/LO/0102). The study registration number was IRAS: 257561. On entry to MINDER participants were assessed using the mini-mental state exam (MMSE) (Folstein, Folstein, and McHugh 1975) and the neuropsychiatric inventory (NPI) (Cummings et al. 1994). Gender and year-of-birth were also recorded for each of the participant and study partner.

### Data from contactless sensors

MINDER includes a range of sensors that are installed in the homes of participants. A key characteristic of the study design is that collection of data is intended to be of minimal burden to the participants. Since our aim was to provide a comprehensive description of behaviour, physiology and how they may be affected by environmental variables we focussed on two specific types of sensors: an under the mattress bedmat (Withings Sleep Analyser (WSA), Withings, France) and an environmental light and temperature multi-sensor device (Develco motion sensor, Develco Products, USA/Denmark). The WSA uses a ballistographic approach to assess when participants are in bed and deduce estimates of physiology and behaviour including sleep parameters, heart rate and breathing rate. Our evaluation of the WSA by comparing its outputs against gold standard polysomnographic measures in older people with and without dementia while attending a sleep laboratory demonstrated that this device provides reliable information on the start and end of major sleep periods, bed entries and bed exits, sleep efficiency as well as heart and breathing rate whereas the estimates of sleep structure (Light Sleep, Deep Sleep, REM sleep) were found to be less reliable (Ravindran et al. 2023a; Ravindran et al. 2023b; Ravindran et al. 2024). Environmental light and temperature multi-sensors were installed in various parts of homes (e.g. kitchen, lounge, bathroom and bedroom). According to the company provided specifications of the sensors they can measure methodology in the laboratory. In our hands the minimum reliably detected level was 4 to 7 lux in light environments with colour temperatures typical of home and office environments (2900K to 5000K). The temperature sensor had an operating range of 0 to 50°C. Whereas the WSA only collected data when participants were in bed, the multi-sensors collect data throughout the 24-hour day. All data were integrated in the MINDER database and extracted from this database using a bespoke software package.

To assess associations between indoor and outdoor environmental light and temperature data from the UK Meteorological office (UKMO) for the analysis period were used. For light, hourly global solar irradiation values (kilojoules / meter square) from the sensor at TW6 2 (West London) were used. Hourly lux values were inferred from the solar irradiation values by multiplying the power in Watts by a value of 105 for the luminous efficiency, representative of average conditions (Littlefair 1985). Hourly temperature values in Celsius were extracted from the dataset MIDAS: UK Hourly Weather Observation Data for the temperature station at postcode SW1A 1 (South West London). The choice of the postcodes for the UKMO was informed by the geographical location of the participants.

### Data selection and curation

All bedmat data and environmental sensor data were plotted for visual inspection. Temperature and light data were considered invalid if there was no change in a 24-hour period (< 0.012% of data). On one night the bedmat recorded exactly the same onset and offset times for sleep across more than 30 participants. This was assumed to be a system failure and the corresponding entries were discarded.

The resulting data set initially comprised a total of 122 participants and approximately 48,000 days. However, some homes did not include data from both the bedmat and data from the environmental multi-sensor, and for some participants data were only available for a small part of the year. Since we were interested in assessing seasonal variation in sleep behaviour and physiology and their association with environmental variables, the year was divided into six two-month periods (Jan-Feb, Mar-Apr etc). Participants were excluded if they did not have simultaneous data from the bedmat, the bedroom light and temperature sensor and lounge light sensor for at least seven 24-hour periods in 5 out of 6 two-month periods.

### Approach to selection of relevant dependent and independent variables

The bedmat reliably assesses bed occupancy. For each minute of a day, the bedmat either does not report any value indicating the bed is unoccupied or reports values that indicate pressure is applied to the bed. In a typical day, there can be multiple short periods when the bed is occupied interspersed with longer periods of bed vacancy. When the bed is occupied, the bedmat stores data at one-minute intervals.

For each minute in a 24-hour period, we determined whether bedmat data were available or not. The simplest measure of duration that was used was to take each 24-hour period and count the number of minutes for which bedmat data were available i.e. the bed was occupied. However, this does not account for the timing and fragmentation of in-bed periods. To construct measures of the timing of bed occupancy, fragmentation and inferred sleep we therefore first defined in-bed bouts where each bout consisted of a contiguous period in which the bedmat registers bed occupancy. In-bed bouts were grouped into in-bed periods, defined as a sequential list of in-bed bouts where the gap between successive in-bed bouts was less than one hour. In-bed periods were classified as “nocturnal” if they had a start time after 18:00 and before 07:00 and as “in-bed periods initiated during the day” otherwise. The longest nocturnal in-bed period was defined as the major nocturnal in-bed period with all other nocturnal in-bed periods classified as minor. (See Supplementary material section S2 for further detail). All in-bed periods had a start time, end time and duration.

Visual inspection of raster plots in which in-bed periods initiated during the day and nocturnal in-bed periods were displayed for each 24 h period and for all participants showed that classification of in-bed periods initiated during the day versus nocturnal in-bed periods was effective in most cases. In most cases, in-bed periods initiated during the day corresponded to conventional views on “naps”.

However, in-bed periods initiated during the day also included periods when people went to bed during the day and then remained in bed for an extended period, as may occur through periods of ill-health. Similarly, some nocturnal in-bed periods extended over multiple days. In our analysis we excluded outliers as defined by a nocturnal time-in-bed period greater than the mean value plus four standard deviations (i.e. those greater than approximately 23 h) and those in which the duration of in-bed periods initiated during the day were longer than the mean value plus three standard deviations (i.e. those greater than approximately 16.5 h). For the 70 participants, this resulted in a final analysis data set consisting of 26,523 24-h periods in which at least one nocturnal time in-bed period occurred.

For the major nocturnal in-bed period multiple further measures related to fragmentation were calculated including: the median duration of in-bed bouts and wake bouts in each in-bed period; the number of bed exits in each in-bed period; the bed exit rate i.e. number of bed exits / duration; the median duration of time out-of-bed in each in-bed period; wake after sleep onset and sleep efficiency. Measures of physiology (mean and standard deviation of heart rate and mean breathing rate during sleep, mean snoring duration) were also calculated. As a marker of the timing of the biological clock we calculated the time of the heart rate minimum during the major nocturnal in-bed period by finding the time of the minimum heart rate for a 1 h moving average. See the Supplementary Material for further details and precise definitions for all metrics used.

In all we selected 25 measures of in-bed occupancy, physiology and timing of the biological clock to analyse in detail. Selection of dependent variables was informed by the reliability of the measurements and their potential relevance to understanding the physiology and behaviour of PLWD. For example, the bedmat reports not only in-bed occupancy but also classifies each in-bed minute as wake, non-REM or REM sleep. We did not consider the measures of non-REM or REM sleep since although they may contain useful information, we have found them not to be reliable measures of non-REM or REM sleep as assessed using gold standard polysomnography in comparable participants.

Dependent variables were sorted into four categories namely 1) Total time in bed 2) Time in bed initiated during the day 3) Time in bed initiated during the night 4) Nocturnal physiology (e.g. heart rate, breathing rate, snoring duration). Categories 2-4 were further sub-divided into measures of number or duration (e.g. number of in-bed periods during the day, number of bed exits, duration of the major nocturnal in-bed period) and measures of timing (e.g. onset time of the major nocturnal in-bed period).

For investigating the daily and seasonal light and temperature environment we constructed hourly averages for light and temperature for each of four rooms (lounge, bedroom, kitchen, bathroom) for each home. Hourly averages took into account the fact that the multi-sensor records changes in light and temperature rather than provides a continuous stream of data (measurements approximately every 20 minutes).

To assess the associations between daily measures of physiology and behaviour, daily summary measures of light and temperature were constructed. These included measures of level (e.g. mean light and mean temperature during the night), hours of bright light (> 500 Lux) and measures of daily change (e.g. difference between the maximum and minimum daily temperature).

### Statistical Methods

We used statistical methods to address the following questions: Do the indoor light and temperature environment vary across homes, position within a home, time of day and seasons? Do indoor light and temperature correlate with outdoor light and temperature? To what extent do measures related to in-bed occupancy, physiology and markers of the biological clock vary between and within participants? Within participants, do measures related to in-bed occupancy, physiology and markers of the biological clock vary with season? After controlling for season, do indoor light and temperature associate with measures of in-bed occupancy, physiology and markers of the biological clock? These questions were addressed using descriptive statistics and linear mixed effect models as appropriate. All computations were done in MATLAB (R2022b) (Inc. 2022) and in most cases cross-checked with SAS (version 9.4) (SAS).

Specifically, to assess how the indoor light and temperature environment vary with time-of-day and position in the house we considered every hour of every day for every participant and considered a fixed effect models in which hour of the day, season (two-month period), room and participant (i.e. home) were entered as categorical variables. To assess whether the indoor light and temperature environment correlates with the external light and temperature, we considered the correlation between daily measures of the outdoor light and temperature environment (geometric mean light level in lux, maximum daily temperature, minimum daily temperature) and the equivalent measures calculated from the indoor multi-sensors averaged across all homes. To illustrate the variation between homes, in addition, we considered the correlation between daily indoor and outdoor environmental measures for each individual home.

To assess seasonal variation of the light and temperature environment within participants we used linear mixed models for daily measures of the light and temperature environment with the fixed effect of season and home (i.e. participant) as a random effect.

To assess the between and within contributions to measures of in-bed occupancy, physiology and markers of the biological clock we calculated intraclass correlation coefficients (ICCs). The ICCs were calculated by first fitting a linear mixed effect model using the MATLAB fitlme function with participant as a random variable, i.e. *y_ij_ =* b*_0_ + b_0j_ +* ɛ_ij_ for observation *y*_ij_ and participant *j*, with the assumptions that *b*_0*j*_ and *ɛ_ij_* are normally distributed, *b_0j_ ∼ N(0,*s*_b_^2^)* and ɛ_ij_ *∼ N(0,*s*^2^)*. Hence,s*_b_^2^* is the between participant variance and *σ*^2^ is the residual variance (within participant variance). The ICC was then calculated from ICC = s*_b_^2^/ (*s*_b_^2^ + σ*^2^).

To assess contributions to within participant variation in measures of in-bed occupancy, physiology and markers of the biological clock we evaluated simple models in which we only controlled for season, age, gender and diagnosis with participant as a random effect, and more complex models in which measures of the indoor environment were also added as co-variates. In the more complex models, we tested whether controlling for season in addition to light and temperature improved the fit of the models. For 22 of the 25 measures that we considered, including season improved the fit of the models according to the Aikake information criterion.

All variables were entered as recorded, i.e. not normalised and all coefficients are therefore in the original units. To facilitate the interpretation of the statistical results we computed the predicted effect of changing the environmental measures (i.e. covariates) over their mean +/-one standard deviation on each measure of interest for an average participant.

## Data availability

The data collected during the current study are available from the corresponding author on reasonable request. The institute intends to store data in a public repository on dementiasplatform.uk in due course.

## Code availability

Code used to analyse the data will be made available on github when the manuscript is accepted.

## Acknowledgments

This work is supported by the UK Dementia Research Institute [award numbers UKDR-7005 DRI-CORE2020-CRT, CF2023/7] through UK DRI Ltd, principally funded by the UK Medical Research Council, and additional funding partner Alzheimer’s Society. We thank the patients and study partners who took part in the study. Surrey and Borders Partnership were also a sponsor of this study. The data were collected as part of the UK Dementia Research Institute Care Research & Technology Centre led by David Sharp (1). A complete list of members is: David Sharp (1), Danielle Wilson (1), Sarah Daniels, Ramin Nilforooshan (2), David Wingfield (3), Matthew Harrison (4), Shlomi Haar (1), Mara Golemme, Payam Barnaghi (1), Paul Freemont (5), Ravi Vaidyanathan (6), Tim Constandinou (7), Gregory Scott (1), Derk-Jan Dijk (8), Pete Lally (11), Paresh Malhotra (1), Louise Robinson (12), Adam Hampshire (1), Michael David (1), Martina Del Giovane (1), Neil Graham (1), Magdalena Kolanko (1), Helen Lai (1), Lucia M Li (1), Mark Crook Rumsey (1), Emma Jane Mallas (1), Alina Irina Serban (1), Eyal Soreq (1), Abidemi Otaiku (1), Megan Parkinson (1), Thomas Parker (1), Success Fabusoro (1), Emily Beal (1), Alan Bannon (7), Danilo Mandic (7), Ziwei Chen (7), Charalambos Hadjipanayi (7), Ghena Hammour (7), Bryan Hsieh (7), Amir Nassibi (7), Adrien Rapeaux (7), Ian Williams (7), Maowen Yin (7), Niro Yogendran (7), Maria Lima (6), Ting Su (6), Melanie Jouaiti (6), Maitreyee Wairagkar (6), Carlos Sebastian Castillo (6), Panipat Wattansiri (6), Thomas Martineau (6), Mayue Shi (6), Tianbo Xu (6), Bo Xiao (6), Alejandro Valdunciel (6), Reneira Seeamber (6), Annika Guez (6), Zehao Liu (6), Saksham Dhawan (6), Nan Fletcher Lloyd (1), Samaneh Kouchaki (9), Alexander Capstick (1), Chloe Walsh (1,2), Louise Rigny (1), Ruxandra Mihai (1), Marirena Bafaloukou (1), Jin Cui (1), Ann-Kathrin Schalkamp (1), Yu Chen (1), Tianyu Cui (1), Nivedita Bijlani (1), Michael Crone (5), Kirsten Jensen (5), Martin Tran (5), Thomas Adam (5), Raphaella Jackson (5), Alexander Webb (5), Anne C Skeldon (10), Kevin Wells (9), Ullrich Bartsch (8), Ciro Della Monica (8), Kiran K G Ravindran (8), Damion Lambert (8), Sara Mohammadi Mahvash (8), Thalia Rodriguez Garcia (8, 10), Victoria L Revell (8), Giuseppe Atzori (8), Lucinda Grainger (8), Hana Hassanin (8), James Woolley (8), Iris Wood-Campar (8), Janetta Rexha (8), Sarmad Al Gawwam (8), Subati Abulikemu (1), Julian Jeyasingh Jacob (1), Nathan Steadman (1), Federico Nardi (1), Cosima Graef (1), Alena Kutuzova (1), Assaf Touboul (1), Nicolas Calvo Peiro (1), Jenna Yun (1), Gaia Frigerio (1), Adela Desowska (1), Anastasia Gailly de Taurines (1), Ruxandra Mihai (1), Nina Moutonnet (1), Sophie Horrocks (4), Brian Quan (4), Mark Woodbridge (1), Anna Joffe (1), Amer Marzuki (1), Ramsheed Abdul Rahim (1), Jessica True (2), Olga Balazikova (2), Nicole Whitethread (2), Matthew Purnell (2), Vaiva Zarombaite (2), Lucy Copps (2), Olivia Knight (2), Gaganpreet Bangar (2), Sumit Dey (2), Chelsea Mukonda (2), Jessica Hine (2), Luke Mallon (2), Saijal Jhala (2), Oliver Sargentoni (2), Amy Alves (2), Mahan Heydari (2), Claire Norman (3), Anesha Patel (3), Ruby Lyall (3), Sanara Raza (3), Naomi Hassim (3), Pippa Kirby (3), John Patterson (3), Mike Law (3), Andy Kenny (3).

(1) Department of Brain Sciences, Faculty of Medicine, Imperial College London, United Kingdom
(2) Surrey and Borders Partnership NHS Foundation Trust, United Kingdom
(3) Hammersmith and Fulham Partnership, United Kingdom
(4) Helix Centre, Faculty of Medicine, Imperial College London, United Kingdom
(5) Department of Infectious Disease, Faculty of Medicine, Imperial College London, United Kingdom
(6) Department of Mechanical Engineering, Faculty of Engineering, Imperial College London, United Kingdom
(7) Department of Electrical and Electronic Engineering, Faculty of Engineering, Imperial College London, United Kingdom
(8) Surrey Sleep Research Centre, School of Biosciences, Faculty of Health and Medical Sciences, University of Surrey, Guildford, United Kingdom.
(9) School of Computer Science and Electronic Engineering, University of Surrey, Guildford, United Kingdom
(10) School of Mathematics & Physics, Faculty of Engineering and Physical Sciences, University of Surrey, Guildford, United Kingdom
(11) Department of Bioengineering, Faculty of Engineering, Imperial College London, United Kingdom
(12) Population Health Sciences Institute, Faculty of Medical Sciences, Newcastle University, United Kingdom

## Author contributions

A.C.S. and T.R. contributed equally to this work as co-authors. Conceptualization: A.C.S, D.J.D. and T.R.; data collection: CR & T; data curation: CR & T, E.S., T.R. & C.W.; formal analyses: A.C.S. and T.R.; visualisation: A.C.S. and T.R.; writing—original draft: A.C.S. and D.J.D.; writing—reviewing: A.C.S, D.J.D., T.R., E.S. & C.W.

## References

1. Analysis, Centre for Environmental Data. 2019. “Met Office MIDAS Open: UK Land Surface Stations Data (1853-current).” In. http://catalogue.ceda.ac.uk/uuid/dbd451271eb04662beade68da43546e1/.

2. Balouch, S., D. A. D. Dijk, J. Rusted, S. S. Skene, N. Tabet, and D. J. Dijk. 2022. ‘Night-to-night variation in sleep associates with day-to-day variation in vigilance, cognition, memory, and behavioral problems in Alzheimer’s disease’, Alzheimers Dement (Amst), 14: e12303.

3. Baniassadi, A., B. Manor, W. Yu, T. Travison, and L. Lipsitz. 2023. ‘Nighttime ambient temperature and sleep in community-dwelling older adults’, Sci Total Environ, 899: 165623.

4. Benca, R., W. J. Herring, R. Khandker, and Z. P. Qureshi. 2022. ‘Burden of Insomnia and Sleep Disturbances and the Impact of Sleep Treatments in Patients with Probable or Possible Alzheimer’s Disease: A Structured Literature Review’, J Alzheimers Dis, 86: 83–109.

5. Benca, R. M., and M. Teodorescu. 2019. ‘Sleep physiology and disorders in aging and dementia’, Handb Clin Neurol, 167: 477–93.

6. Biller, A. M., N. Fatima, C. Hamberger, L. Hainke, V. Plankl, A. Nadeem, A. Kramer, M. Hecht, and M. Spitschan. 2024. ‘The Ecology of Human Sleep (EcoSleep) Cohort Study: Protocol for a longitudinal repeated measurement burst design study to assess the relationship between sleep determinants and outcomes under real-world conditions across time of year’, J Sleep Res: e14225.

7. Bodenstein, C., M. Gosak, S. Schuster, M. Marhl, and M. Perc. 2012. ‘Modeling the seasonal adaptation of circadian clocks by changes in the network structure of the suprachiasmatic nucleus’, PLoS Comput Biol, 8: e1002697.

8. Boiko, A., N. Martinez Madrid, and R. Seepold. 2023. ‘Contactless Technologies, Sensors, and Systems for Cardiac and Respiratory Measurement during Sleep: A Systematic Review’, Sensors (Basel), 23.

9. Bowman, C., Y. Huang, O. J. Walch, Y. Fang, E. Frank, J. Tyler, C. Mayer, C. Stockbridge, C. Goldstein, S. Sen, and D. B. Forger. 2021. ‘A method for characterizing daily physiology from widely used wearables’, Cell Rep Methods, 1.

10. Brown, T. M., G. C. Brainard, C. Cajochen, C. A. Czeisler, J. P. Hanifin, S. W. Lockley, R. J. Lucas, M. Munch, J. B. O’Hagan, S. N. Peirson, L. L. A. Price, T. Roenneberg, L. J. M. Schlangen, D. J. Skene, M. Spitschan, C. Vetter, P. C. Zee, and K. P. Wright, Jr. 2022. ‘Recommendations for daytime, evening, and nighttime indoor light exposure to best support physiology, sleep, and wakefulness in healthy adults’, PLoS Biol, 20: e3001571.

11. Burns, A. C., R. Saxena, C. Vetter, A. J. K. Phillips, J. M. Lane, and S. W. Cain. 2021. ‘Time spent in outdoor light is associated with mood, sleep, and circadian rhythm-related outcomes: A cross-sectional and longitudinal study in over 400,000 UK Biobank participants’, J Affect Disord, 295: 347–52.

12. Butler, M. P., C. Smales, H. Wu, M. V. Hussain, Y. A. Mohamed, M. Morimoto, and S. A. Shea. 2015. ‘The Circadian System Contributes to Apnea Lengthening across the Night in Obstructive Sleep Apnea’, Sleep, 38: 1793–801.

13. Casagrande, M., G. Forte, F. Favieri, and I. Corbo. 2022. ‘Sleep Quality and Aging: A Systematic Review on Healthy Older People, Mild Cognitive Impairment and Alzheimer’s Disease’, Int J Environ Res Public Health, 19.

14. Cederroth, C. R., U. Albrecht, J. Bass, S. A. Brown, J. DyhrÖeld-Johnsen, F. Gachon, C. B. Green, M. H. Hastings, C. Helfrich-Forster, J. B. Hogenesch, F. Levi, A. Loudon, G. B. Lundkvist, J. H. Meijer, M. Rosbash, J. S. Takahashi, M. Young, and B. Canlon. 2019. ‘Medicine in the Fourth Dimension’, Cell Metab, 30: 238–50.

15. Chevance, G., K. Minor, C. Vielma, E. Campi, C. O’Callaghan-Gordo, X. Basagana, J. Ballester, and P. Bernard. 2024. ‘A systematic review of ambient heat and sleep in a warming climate’, Sleep Med Rev, 75: 101915.

16. Cummings, J. L., M. Mega, K. Gray, S. Rosenberg-Thompson, D. A. Carusi, and J. Gornbein. 1994. ‘The Neuropsychiatric Inventory: comprehensive assessment of psychopathology in dementia’, Neurology, 44: 2308–14.

17. Dijk, D. J., and C. A. Czeisler. 1995. ‘Contribution of the circadian pacemaker and the sleep homeostat to sleep propensity, sleep structure, electroencephalographic slow waves, and sleep spindle activity in humans’, J Neurosci, 15: 3526–38.

18. Falgas, N., C. M. Walsh, L. Yack, A. J. Simon, I. E. Allen, J. H. Kramer, H. J. Rosen, R. Joie, G. Rabinovici, B. Miller, S. Spina, W. W. Seeley, K. Ranasinghe, K. Vossel, T. C. Neylan, and L. T. Grinberg. 2023. ‘Alzheimer’s disease phenotypes show different sleep architecture’, Alzheimers Dement, 19: 3272–82.

19. Folstein, M. F., S. E. Folstein, and P. R. McHugh. 1975. ‘“Mini-mental state”. A practical method for grading the cognitive state of patients for the clinician’, J Psychiatr Res, 12: 189–98.

20. Gaggioni, G., P. Maquet, C. Schmidt, D. J. Dijk, and G. Vandewalle. 2014. ‘Neuroimaging, cognition, light and circadian rhythms’, Front Syst Neurosci, 8: 126.

21. Harding, E. C., N. P. Franks, and W. Wisden. 2020. ‘Sleep and thermoregulation’, Curr Opin Physiol, 15: 7–13.

22. Hayter, E. A., S. M. T. Wehrens, H. P. A. Van Dongen, A. Stangherlin, S. Gaddameedhi, E. Crooks, N. J. Barron, L. A. Venetucci, J. S. O’Neill, T. M. Brown, D. J. Skene, A. W. Trafford, and D. A. Bechtold. 2021. ‘Distinct circadian mechanisms govern cardiac rhythms and susceptibility to arrhythmia’, Nat Commun, 12: 2472.

23. Inc., The MathWorks. 2022. “MATLAB.” In.: Natick, Massachusetts: The MathWorks Inc.

24. Klepeis, N. E., W. C. Nelson, W. R. Ott, J. P. Robinson, A. M. Tsang, P. Switzer, J. V. Behar, S. C. Hern, and W. H. Engelmann. 2001. ‘The National Human Activity Pattern Survey (NHAPS): a resource for assessing exposure to environmental pollutants’, J Expo Anal Environ Epidemiol, 11: 231–52.

25. Klerman, E. B., and D. J. Dijk. 2005. ‘Interindividual variation in sleep duration and its association with sleep debt in young adults’, Sleep, 28: 1253–9.

26. Koren, T., E. Fisher, L. Webster, G. Livingston, and P. Rapaport. 2023. ‘Prevalence of sleep disturbances in people with dementia living in the community: A systematic review and meta-analysis’, Ageing Res Rev, 83: 101782.

27. Lan, L., K. Tsuzuki, Y. F. Liu, and Z. W. Lian. 2017. ‘Thermal environment and sleep quality: A review’, Energy and Buildings, 149: 101–13.

28. LeGates, T. A., D. C. Fernandez, and S. Hattar. 2014. ‘Light as a central modulator of circadian rhythms, sleep and affect’, Nat Rev Neurosci, 15: 443–54.

29. Li, L., T. Nakamura, J. Hayano, and Y. Yamamoto. 2021. ‘Seasonal Sleep Variations and Their Association With Meteorological Factors: A Japanese Population Study Using Large-Scale Body Acceleration Data’, Front Digit Health, 3: 677043.

30. Lim, A. S. P., C. Gaiteri, L. Yu, S. Sohail, W. Swardfager, S. Tasaki, J. A. Schneider, C. Paquet, D. T. Stuss, M. Masellis, S. E. Black, J. Hugon, A. S. Buchman, L. L. Barnes, D. A. Bennett, and P. L. De Jager. 2018. ‘Seasonal plasticity of cognition and related biological measures in adults with and without Alzheimer disease: Analysis of multiple cohorts’, PLoS Med, 15: e1002647.

31. Lincoln, G. 2019. ‘A brief history of circannual time’, J Neuroendocrinol, 31: e12694.

32. Littlefair, Paul J. 1985. ‘The luminous efficacy of daylight: a review’, Lighting Research & Technology, 17: 162–82.

33. Lomas, K., K. Morgan, V. Haines, I. Hartescu, A. Beizaee, J. Barnes, Z. Zambelli, M. Ravikumar, and V. Rossi. 2024. ‘Homes Heat Health protocol: an observational cohort study measuring the effect of summer temperatures on sleep quality’, BMJ Open, 14: e086797.

34. Lomas, Kevin J, and Matthew Li. 2023. ‘An overheating criterion for bedrooms in temperate climates: Derivation and application’, Building Services Engineering Research and Technology, 44: 485–517.

35. Lucas, R. J., S. N. Peirson, D. M. Berson, T. M. Brown, H. M. Cooper, C. A. Czeisler, M. G. Figueiro, P. D. Gamlin, S. W. Lockley, J. B. O’Hagan, L. L. Price, I. Provencio, D. J. Skene, and G. C. Brainard. 2014. ‘Measuring and using light in the melanopsin age’, Trends Neurosci, 37: 1–9.

36. Lysen, T. S., A. I. Luik, M. K. Ikram, H. Tiemeier, and M. A. Ikram. 2020. ‘Actigraphy-estimated sleep and 24-hour activity rhythms and the risk of dementia’, Alzheimers Dement, 16: 1259–67.

37. Meyer, N., A. G. Harvey, S. W. Lockley, and D. J. Dijk. 2022. ‘Circadian rhythms and disorders of the timing of sleep’, Lancet, 400: 1061–78.

38. Meyer, N., R. Lok, C. Schmidt, S. D. Kyle, C. A. McClung, C. Cajochen, Fajl Scheer, M. W. Jones, and S. L. Chellappa. 2024. ‘The sleep-circadian interface: A window into mental disorders’, Proc Natl Acad Sci U S A, 121: e2214756121.

39. Mooldijk, S. S., S. Licher, M. W. Vernooij, M. K. Ikram, and M. A. Ikram. 2022. ‘Seasonality of cognitive function in the general population: the Rotterdam Study’, Geroscience, 44: 281–91.

40. Natarajan, A., K. Gleichauf, M. Khalid, C. Heneghan, and L. D. Schneider. 2025. ‘Circadian rhythm of heart rate and activity: A cross-sectional study’, Chronobiol Int, 42: 108–21.

41. Nguyen, J. L., J. Schwartz, and D. W. Dockery. 2014. ‘The relationship between indoor and outdoor temperature, apparent temperature, relative humidity, and absolute humidity’, Indoor Air, 24: 103–12.

42. Obradovich, N., R. Migliorini, S. C. Mednick, and J. H. Fowler. 2017. ‘Nighttime temperature and human sleep loss in a changing climate’, Sci Adv, 3: e1601555.

43. Ravindran, K. K. G., C. Della Monica, G. Atzori, D. Lambert, H. Hassanin, V. Revell, and D. J. Dijk. Ravindran, K. K. G., C. Della Monica, G. Atzori, D. Lambert, H. Hassanin, V. Revell, and D. J. Dijk. ‘Contactless and longitudinal monitoring of nocturnal sleep and daytime naps in older men and women: a digital health technology evaluation study’, Sleep, 46.

44. ———. 2024. ‘Reliable Contactless Monitoring of Heart Rate, Breathing Rate, and Breathing Disturbance During Sleep in Aging: Digital Health Technology Evaluation Study’, JMIR Mhealth Uhealth, 12: e53643.

45. Ravindran, K. K.G., C. Della Monica, G. Atzori, D. Lambert, H. Hassanin, V. Revell, and D. J. Dijk. 2023b. ‘Three Contactless Sleep Technologies Compared With Actigraphy and Polysomnography in a Heterogeneous Group of Older Men and Women in a Model of Mild Sleep Disturbance: Sleep Laboratory Study’, JMIR Mhealth Uhealth, 11: e46338.

46. Rigny, L., N. Fletcher-Lloyd, A. Capstick, R. Nilforooshan, and P. Barnaghi. 2024. ‘Assessment of sleep patterns in dementia and general population cohorts using passive in-home monitoring technologies’, Commun Med (Lond), 4: 222.

47. Roenneberg, T. 2004. ‘The decline in human seasonality’, J Biol Rhythms, 19: 193-5; discussion 96-7.

48. Roenneberg, T., K. V. Allebrandt, M. Merrow, and C. Vetter. 2012. ‘Social jetlag and obesity’, Curr Biol, 22: 939-43.

49. SAS.

50. Schlangen, L. J. M., and L. L. A. Price. 2021. ‘The Lighting Environment, Its Metrology, and Non-visual Responses’, Front Neurol, 12: 624861.

51. Shen, Y., Q. K. Lv, W. Y. Xie, S. Y. Gong, S. Zhuang, J. Y. Liu, C. J. Mao, and C. F. Liu. 2023. ‘Circadian disruption and sleep disorders in neurodegeneration’, Transl Neurodegener, 12: 8.

52. Shochat, T., N. Santhi, P. Herer, S. A. Flavell, A. C. Skeldon, and D. J. Dijk. 2019. ‘Sleep Timing in Late Autumn and Late Spring Associates With Light Exposure Rather Than Sun Time in College Students’, Front Neurosci, 13: 882.

53. Skeldon, A. C., and D. J. Dijk. 2021. ‘Weekly and seasonal variation in the circadian melatonin rhythm in humans: Entrained to local clock time, social time, light exposure or sun time?’, J Pineal Res, 71: e12746.

54. Skeldon, A. C., A. J. Phillips, and D. J. Dijk. 2017. ‘The effects of self-selected light-dark cycles and social constraints on human sleep and circadian timing: a modeling approach’, Sci Rep, 7: 45158.

55. Skorucak, J., E. L. Arbon, D. J. Dijk, and P. Achermann. 2018. ‘Response to chronic sleep restriction, extension, and subsequent total sleep deprivation in humans: adaptation or preserved sleep homeostasis?’, Sleep, 41.

56. Spengler, C. M., C. A. Czeisler, and S. A. Shea. 2000. ‘An endogenous circadian rhythm of respiratory control in humans’, J Physiol, 526 Pt 3: 683–94.

57. Suzuki, M., T. Taniguchi, R. Furihata, K. Yoshita, Y. Arai, N. Yoshiike, and M. Uchiyama. 2019. ‘Seasonal changes in sleep duration and sleep problems: A prospective study in Japanese community residents’, PLoS One, 14: e0215345.

58. Thorne, H. C., K. H. Jones, S. P. Peters, S. N. Archer, and D. J. Dijk. 2009. ‘Daily and seasonal variation in the spectral composition of light exposure in humans’, Chronobiol Int, 26: 854–66.

59. Titova, O. E., E. Lindberg, S. Elmstahl, L. Lind, and C. Benedict. 2022. ‘Seasonal variations in sleep duration and sleep complaints: A Swedish cohort study in middle-aged and older individuals’, J Sleep Res, 31: e13453.

60. Vandewalle, G., B. Middleton, S. M. Rajaratnam, B. M. Stone, B. Thorleifsdottir, J. Arendt, and D. J. Dijk. 2007. ‘Robust circadian rhythm in heart rate and its variability: influence of exogenous melatonin and photoperiod’, J Sleep Res, 16: 148–55.

61. Wang, C., and D. M. Holtzman. 2020. ‘Bidirectional relationship between sleep and Alzheimer’s disease: role of amyloid, tau, and other factors’, Neuropsychopharmacology, 45: 104–20.

62. Wilckens, K. A., R. F. Habte, Y. Dong, M. E. Stepan, K. M. Dessa, A. B. Whitehead, C. W. Peng, M. E. Fletcher, and D. J. Buysse. 2024. ‘A pilot time-in-bed restriction intervention behaviorally enhances slow-wave activity in older adults’, Front Sleep, 2.

63. Windred, D. P., A. C. Burns, J. M. Lane, P. Olivier, M. K. Rutter, R. Saxena, A. J. K. Phillips, and S. W. Cain. 2024. ‘Brighter nights and darker days predict higher mortality risk: A prospective analysis of personal light exposure in >88,000 individuals’, Proc Natl Acad Sci U S A, 121: e2405924121.

64. Winsky-Sommerer, R., P. de Oliveira, S. Loomis, K. Wafford, D. J. Dijk, and G. Gilmour. 2019. ‘Disturbances of sleep quality, timing and structure and their relationship with other neuropsychiatric symptoms in Alzheimer’s disease and schizophrenia: Insights from studies in patient populations and animal models’, Neurosci Biobehav Rev, 97: 112–37.

65. Wyse, C., G. O’Malley, A. N. Coogan, S. McConkey, and D. J. Smith. 2021. ‘Seasonal and daytime variation in multiple immune parameters in humans: Evidence from 329,261 participants of the UK Biobank cohort’, iScience, 24: 102255.

66. Yu, Z., N. Hu, Y. Du, H. Wang, L. Pu, X. Zhang, D. Pan, X. He, and J. Li. 2022. ‘Association of outdoor artificial light at night with mental health among China adults: a prospective ecology study’, Environ Sci Pollut Res Int, 29: 82286–96.

67. Zhai, H., Y. Yan, S. He, P. Zhao, and B. Zhang. 2023. ‘Evaluation of the Accuracy of Contactless Consumer Sleep-Tracking Devices Application in Human Experiment: A Systematic Review and Meta-Analysis’, Sensors (Basel), 23.

68. Zhang, R., and N. D. Volkow. 2023. ‘Seasonality of brain function: role in psychiatric disorders’, Transl Psychiatry, 13: 65.

69. Zhang, Y., A. A. Folarin, S. Sun, N. Cummins, Y. Ranjan, Z. Rashid, C. Stewart, P. Conde, H. Sankesara, P. Laiou, F. Matcham, K. M. White, C. Oetzmann, F. Lamers, S. Siddi, S. Simblett, S. Vairavan, I. Myin-Germeys, D. C. Mohr, T. Wykes, J. M. Haro, P. Annas, B. W. Penninx, V. A. Narayan, M. Hotopf, R. J. Dobson, and Radar-Cns consortium. 2024. ‘Longitudinal Assessment of Seasonal Impacts and Depression Associations on Circadian Rhythm Using Multimodal Wearable Sensing: Retrospective Analysis’, J Med Internet Res, 26: e55302.

70. Zolfaghari, S., M. Cyr, A. Pelletier, and R. B. Postuma. 2023. ‘Effects of Season and Daylight Saving Time Shifts on Sleep Symptoms: Canadian Longitudinal Study on Aging’, Neurology, 101: e74–e82.

