## Supplementary Material for "Seasonal and daily variation in indoor light and temperature associate with sleep disturbance in dementia"

S4 **Seasonal variation of in-bed duration, timing and fragmentation, physiology.** The data shown are the Least Squares (LS) means from a linear mixed effect model which considers the seasonal variation for each measure controlling for gender, age and diagnosis with participant as a random effect (see Supplementary Table S6). Eight of the 25 measures are also shown in the Fig. 3 of the main text. All measures are included here for ease of visual comparison. Times are reported in both local clock time and GMT. The effect of reporting times in GMT is to subtract one hour to the times in May/Jun, Jul/Aug and for some days in Mar/Apr and Sep/Oct which are the periods which cover the GMT to daylight saving time transitions. In, for example, the onset times of the major nocturnal in-bed period, it can clearly be seen how the subtraction of one hour for part of the year induces a large seasonal change which is not present in the data reported in local clock time. . . . . . . . . . . . . . . . . . . . . . . . . . . . . . . . . . . . . . . . . . . . 11

S5 **Individual variation in total time in-bed.** Time courses of the total time in-bed for each participants. Each data point is a single day. The magenta lines show the two-month moving averages. . . . . . . . . . . . . . . . . . . . . . . . . . . . . . . . . . . . . . . 12

S6 **Desynchrony due to a maladaptive light environment?** a) Raster plot for the heart rate for participant "seasonal", replicated from Fig. 1 (b). Recording of heart rate during sleep for one night showing how the heart rate dips to a minimum. Superimposed on the raw data is a 60 minute moving average, with the circular marker indicating the minimum. (c) Mean daily light levels in the lounge for participant "seasonal". (d) Raster plots of sleep predictions. In the left hand panel, it is assumed that light is not gated by sleep. In the right hand panel, it is assumed that the measured light is gated by sleep. . . . . . . . . . . . . 20

S4 **Descriptive statistics for the measures of sleep behaviour and physiology and the light and temperature environment** Descriptive statistics describing the distribution of the $n = 70$ participant mean values for the 25 different measures of sleep behaviour and physiology and 4 measures of the light and temperature environment. The intraclass correlation coefficient for each measure is also included. For all measures, the reported mean is the mean of the participant mean values, i.e. from the approximately 26,000 days of data, the individual participant mean values were first calculated. Then the mean of the participant values was found. . . . . . . . . . . . . . . . . . . . . . . . . . . . . . 14

S5 **Correlation between bedmat variables and measures of age, cognition, behaviour and environmental light and temperature.** Correlation coefficients (Spearman's rho) and p values were calculated as the correlation between participant mean values (n=70). 15

S6 **Seasonal effects on the duration, timing and fragmentation of bed occupancy and physiology.** Results from a linear mixed effect model for the seasonal variation of the 25 measures of bed occupancy, physiology and a marker of the biological clock controlling for gender, age and diagnosis. One of the participants did not have a diagnosis, so only 69 of the 70 participants were included in this analysis. . . . . . . . . . . . . . . . . . . . . 16

S7 **Effects of light, temperature and time-in-study on the duration, timing and fragmentation of bed occupancy and on physiology.** Linear mixed effect model results for the 25 measures of bed occupancy and physiology, extending the model reported in Table S6 to include environmental variables. Only the results for the additional variables (hours of bright light, mean nocturnal temperature, temperature amplitude and time-in-study) are shown. All other effects are similar to those reported in Supplementary Table S6). One of the participant's did not have a diagnosis, so only 69 of the 70 participants were included in this analysis. . . . . . . . . . . . . . . . . . . . . . . . . . . . . . . . . . . . . . . 17

S8 **Correlation between the seasonal change in duration, onset and offset of the major time-in-bed period with age, MMSE and NPI and the seasonal change in environmental light and temperature.** Correlation coefficients (Spearman's rho) and p values were calculated as the correlation between participant mean values (n=70). The seasonal change for each participant was calculated as the mean winter (Jan-Feb) value minus the mean summer (Jul-Aug) value. . . . . . . . . . . . . . . . . . . . . . . . . . . . . 18

### S1 Demographics

| | Number of participants | Age (+/-SD) (y) | MMSE (+/-SD) | Total NPI score (+/-SD) | NPI sleep score (+/-SD) | ADAS Cog (+/-SD) | Collection period (+/-SD) (d) | Days of data (+/-SD) (d) |
|---|---|---|---|---|---|---|---|---|
| **All** | 70 | 79 (+/-9) | 22 (+/-6) | 12 (+/-10) | 1.3 (+/-2.4) | 36 (+/-16) | 464 (+/-169) | 379 (+/-149) |
| **Men** | 44 | 78 (+/-9) | 23 (+/-5) | 11 (+/-10) | 1.0 (+/-2.0) | 37 (+/-16) | 480 (+/-178) | 393 (+/-154) |
| **Women** | 26 | 81 (+/-9) | 20 (+/-8) | 13 (+/-10) | 1.8 (+/-2.8) | 33 (+/-15) | 435 (+/-153) | 355 (+/-139) |
| **Alzheimer's diagnosis** | 48 | 80 (+/-8) | 21 (+/-7) | 11 (+/-10) | 0.9 (+/-2.1) | 38 (+/-14) | 447 (+/-157) | 368 (+/-135) |
| **Other diagnosis** | 21 | 78 (+/-11) | 22 (+/-5) | 14 (+/-11) | 2.0 (+/-2.8) | 30 (+/-17) | 511 (+/-191) | 410 (+/-179) |

Table S1: **Participant demographics.**

#### S2 Definitions

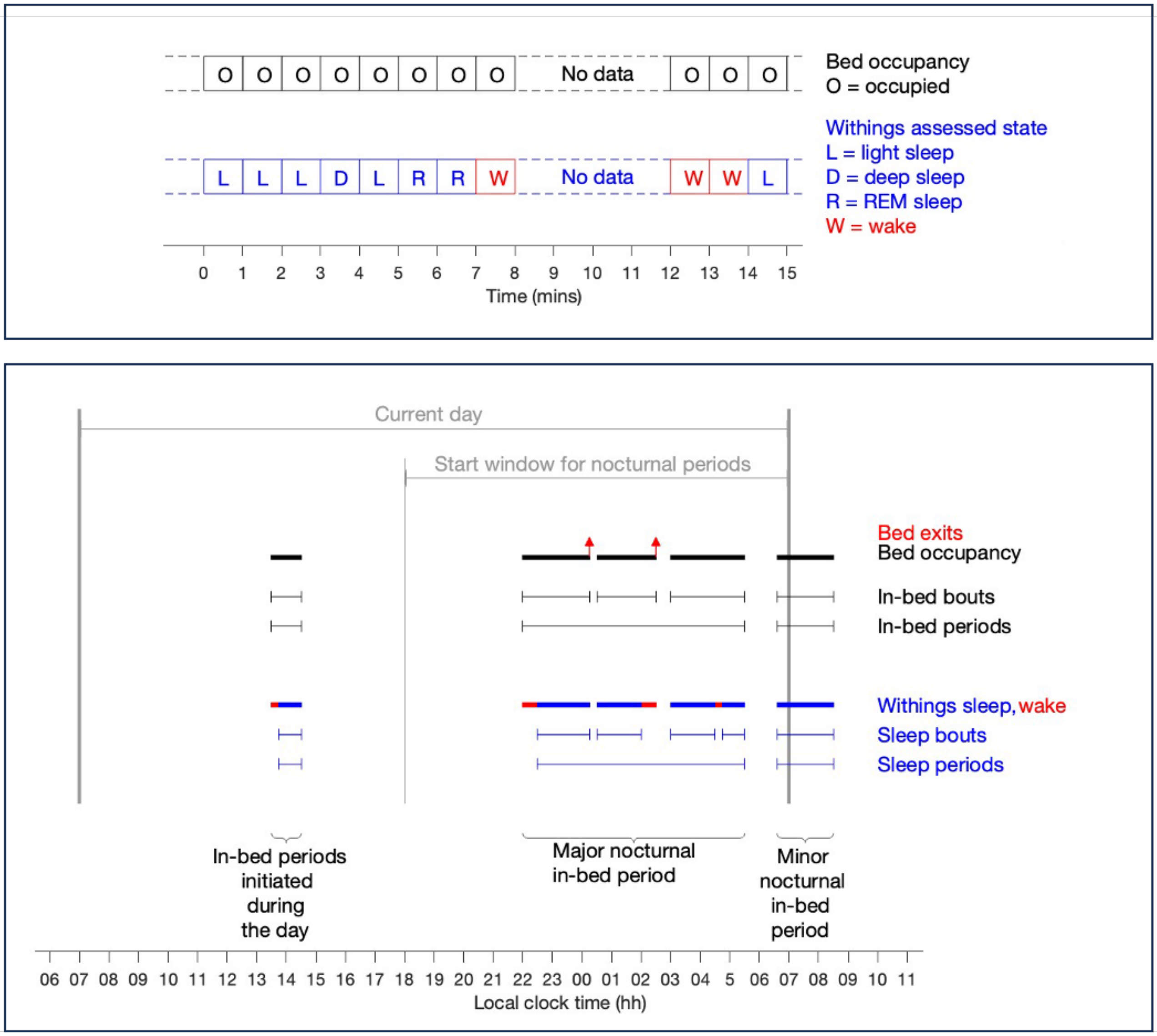

Figure S1: **Derivation of in-bed and sleep metrics from the bedmat.** Graphical depiction of the process for defining metrics from the Withing's sleep analyser (WSA) bedmat. The top panel illustrates the nature of the minute-to-minute data streamed from the bedmat. The lower panel describes how the minute-to-minute data is used to define in-bed and sleep periods and associated metrics.

**Definitions**

**Total time in-bed (h)** Total time that the Withings sleep analyser (WSA) registers that someone is in bed between 07:00 and 07:00.

**In-bed bout** A contiguous sequence of minutes in which the WSA registers that an individual is in bed. An in-bed bout has a start time, an end time and a duration (h).

**Out-of-bed bouts** An unbroken period of time in which the WSA registers no information.

**Sleep bout** A contiguous sequence of minutes in which the WSA registers that an individual is in one of deep, light or REM sleep. A sleep bout has a start time, an end time and a duration (h).

**In-bed period** A sequential group of in-bed bouts where the gap between successive in-bed bouts is less than one hour.

**Sleep period** A sequential group of sleep bouts where the gap between successive sleep bouts is less than one hour.

**Nocturnal in-bed period** An in-bed period where the start time of the first in-bed bout is after 18:00 and before 07:00 and at least 2 minutes are recorded as sleep.

**Nocturnal sleep period** A sleep period where the start time of the first sleep bout is after 18:00 and before 07:00 and the period is at least 2 minutes long.

**Major nocturnal in-bed period** The longest nocturnal in-bed period.

**Major nocturnal sleep period** The longest nocturnal sleep period.

**Minor nocturnal in-bed periods** All nocturnal in-bed periods other than the major nocturnal in-bed period.

**Minor nocturnal sleep periods** All nocturnal sleep periods other than the major nocturnal sleep period.

**Current day** The day leading up to the nocturnal in-bed period. For daily measures (e.g. the total time in-bed, the current day starts at 07:00.

**Definitions related to in-bed periods initiated during the day**

**In-bed period initiated during the day** An in-bed period with the start time of the first in-bed bout in the period after 07:00 and before 18:00 and at least 2 minutes are recorded as sleep.

**Sleep period initiated during the day** A sleep period with the start time of the first in-bed bout in the period after 07:00 and before 18:00 and the period is at least 2 minutes long.

**Total duration of in-bed periods initiated during the day on the current day (h)** The sum of the durations of all the in-bed periods that start during the day on the current day.

**Definitions related to the major nocturnal period**

**Duration of the major nocturnal in-bed period (h)** The period of time from the start of the major nocturnal in-bed period to the end of the major nocturnal in-bed period.

**Onset time of the major nocturnal in-bed period** The start time of the major nocturnal in-bed period.

**Offset time of the major nocturnal time-in-bed period** The end time of the major nocturnal in-bed period.

**Definitions related to fragmentation**

**Number of bed exits** Number of out-of-bed bouts during the major nocturnal time-in-bed period.

**Bed exit rate (exits/h)** Number of 'bed exits' / Major nocturnal in-bed period duration.

**Median duration of bed exits (h)** The median duration of out-of-bed bouts during the major nocturnal in-bed period.

**Median duration of sleep bouts (h)** The median duration of sleep bouts during the major nocturnal in-bed period.

**Median duration of wake bouts (h)** The median duration of wake bouts during the major nocturnal in-bed period.

**Wake after sleep onset (h)** The sum of the WSA wake state duration and the out-of-bed duration during the major nocturnal sleep period.

**Sleep efficiency (%)** 100 $\times$ The nocturnal sleep duration / duration of the major nocturnal in-bed period.

**Definitions related to physiology**

**Mean breathing rate (breaths/min)** Mean breathing rate during the major nocturnal sleep period for minutes that the WSA registers that an individual is asleep.

**Mean heart rate (beats/min)** Mean heart rate recorded during the major nocturnal sleep period for minutes that the WSA registers than an individual is asleep.

**Std dev. heart rate (beats/min)** Standard deviation of the heart rate values recorded during the major nocturnal sleep period for minutes that the WSA registers than an individual is asleep.

**Definitions related to the environment**

**Mean light intensity (lux)** The mean light intensity recorded by the multi-sensor between 07:00 on the current day and 07:00 on the next day.

**Geometric mean light intensity (lux)** The geometric mean light intensity recorded by the multi-sensor between 07:00 on the current day and 07:00 on the next day.

**Hours of bright light (h)** The amount of time that the multi-sensor records light levels of 500 or more lux on the current day.

**Mean daily temperature (°C)** The mean temperature recorded by the multi-sensor between 07:00 on the current day and 07:00 on the next day.

**Maximum daily temperature (°C)** The maximum temperature recorded by the multi-sensor between 07:00 on the current day and 07:00 on the next day.

**Minimum daily temperature (°C)** The minimum temperature recorded by the multi-sensor between 07:00 on the current day and 07:00 on the next day.

**Mean nocturnal temperature (°C)** The mean temperature recorded by the multi-sensor during the major nocturnal in-bed period.

**Temperature amplitude (°C)** The difference between the maximum and minimum value of the temperature on each 24-hour period from 07:00 on the current day to 07:00 on the next day.

### S3 Further figures

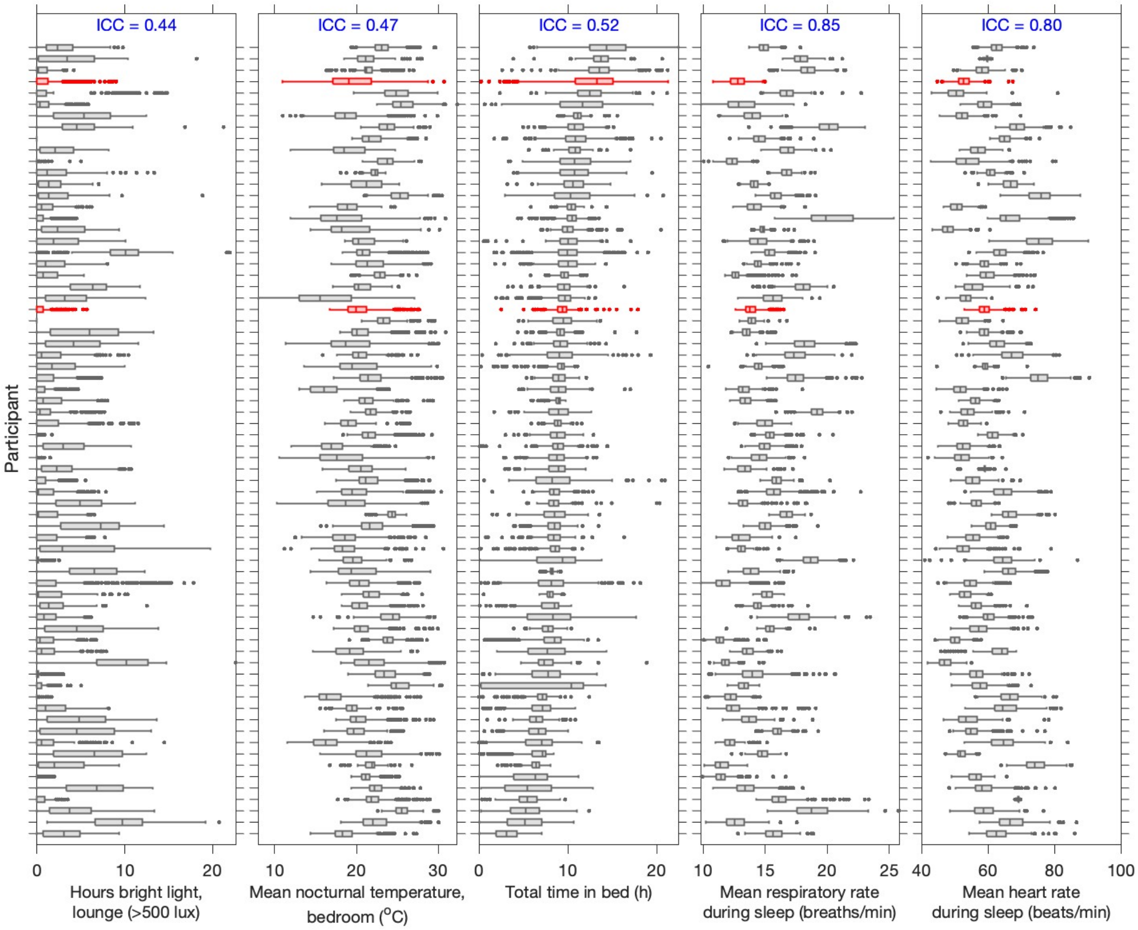

Figure S2: **Between and within participant variation in indoor light and temperature environment, bed occupancy, respiratory rate and heart rate.** Box plots for each participant showing the between and within participant variation in a few example metrics that quantify aspects of the indoor daytime light environment, the nighttime temperature environment, time in bed, respiratory rate and heart rate.

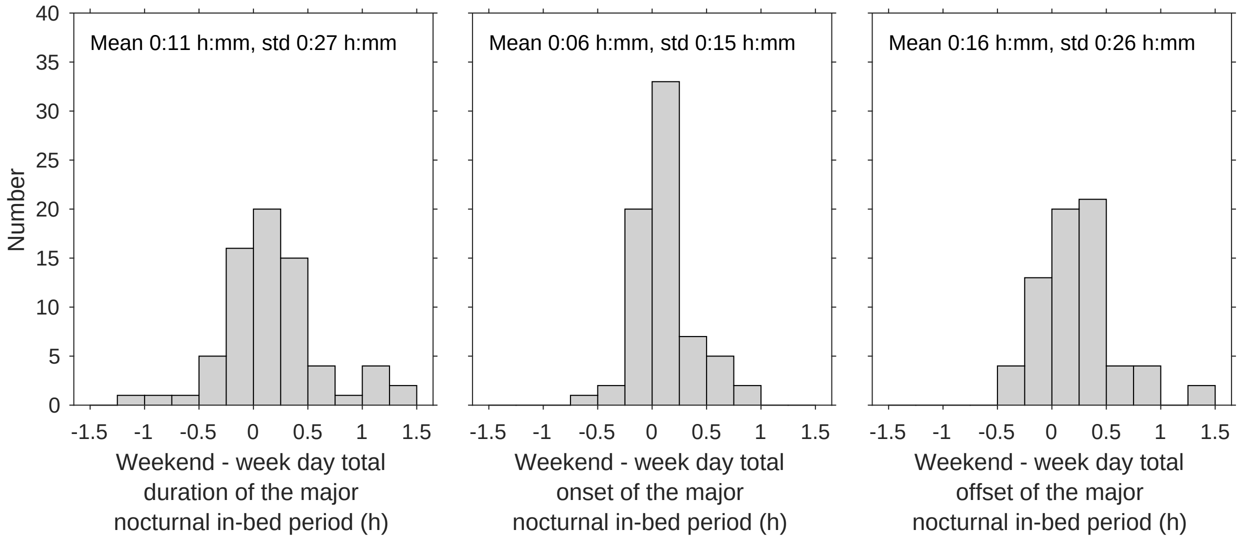

Figure S3: **Histograms of the difference between the mean weekend and week day duration, onset and offset of the major nocturnal in-bed period per participant (n=70).**

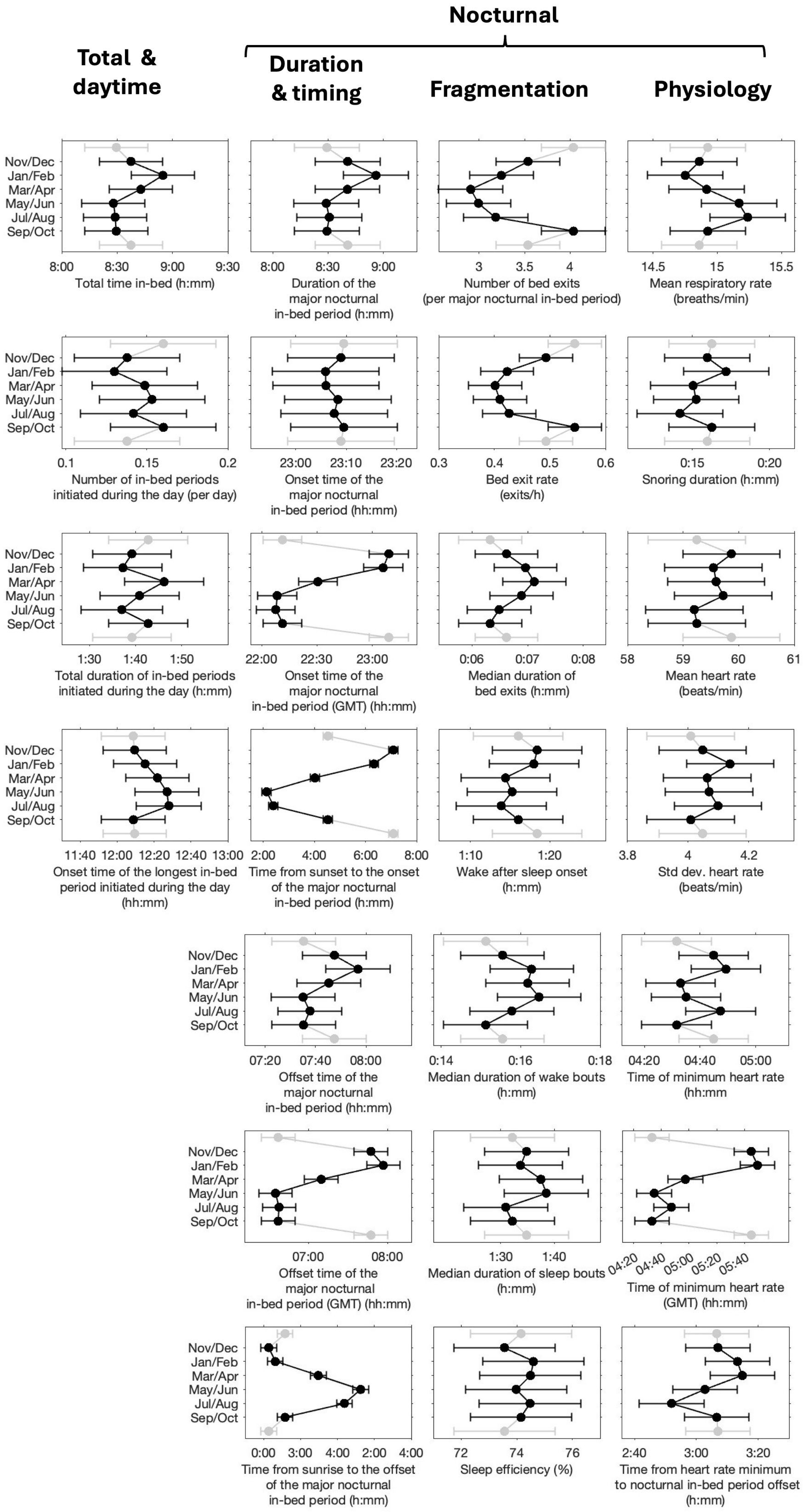

Figure S4: **Seasonal variation of in-bed duration, timing and fragmentation, physiology.** The data shown are the Least Squares (LS) means from a linear mixed effect model which considers the seasonal variation for each measure controlling for gender, age and diagnosis with participant as a random effect (see Supplementary Table S6). Eight of the 25 measures are also shown in the Fig. 3 of the main text. All measures are included here for ease of visual comparison. Times are reported in both local clock time and GMT. The effect of reporting times in GMT is to subtract one hour to the times in May/Jun, Jul/Aug and for some days in Mar/Apr and Sep/Oct which are the periods which cover the GMT to daylight saving time transitions. In, for example, the onset times of the major nocturnal in-bed period, it can clearly be seen how the subtraction of one hour for part of the year induces a large seasonal change which is not present in the data reported in local clock time.

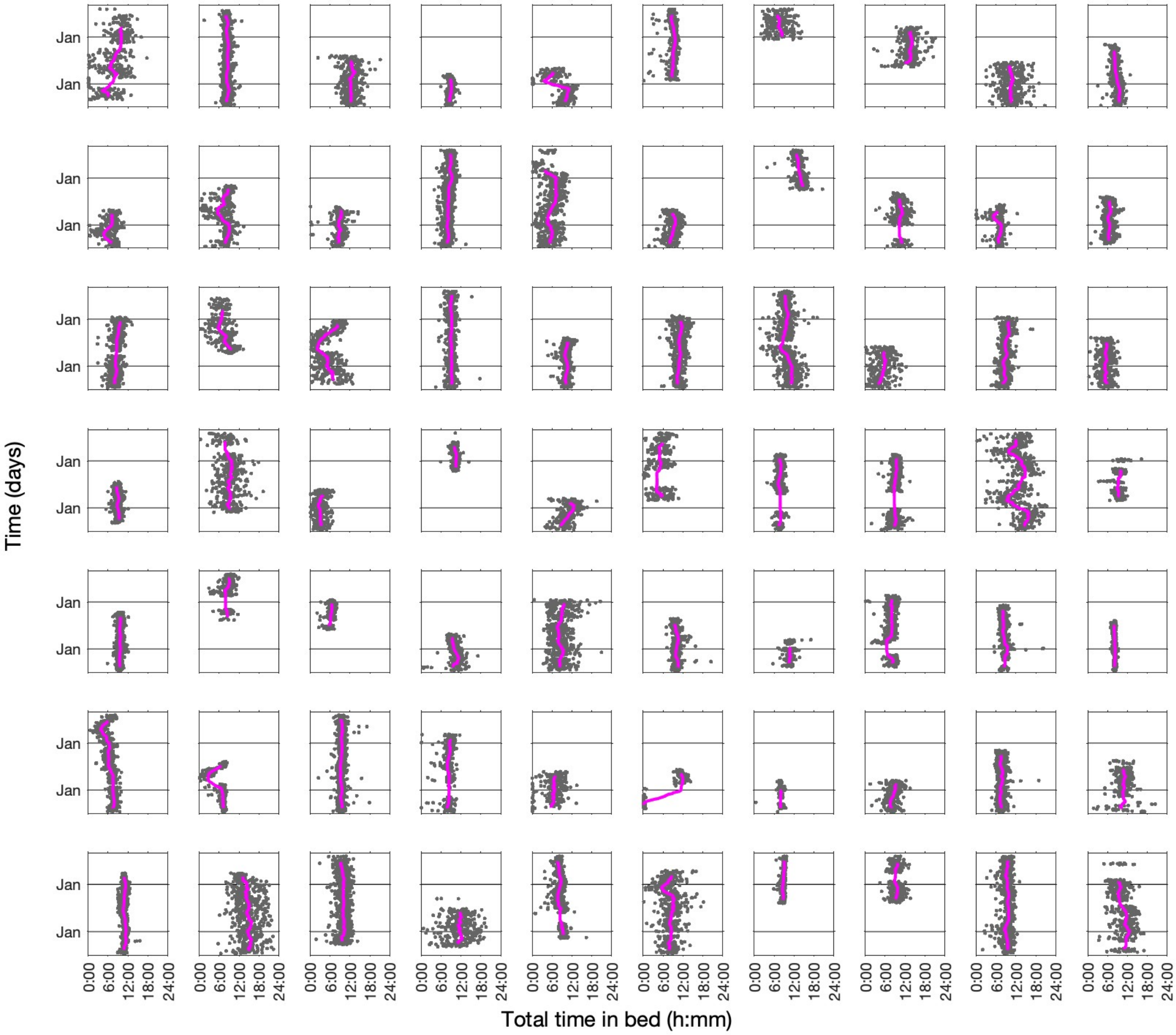

Figure S5: **Individual variation in total time in-bed.** Time courses of the total time in-bed for each participants. Each data point is a single day. The magenta lines show the two-month moving averages.

#### S4 Tables summarising statistical results

| | Two-month period | | | | Hour of the day | | | | Room | | | | Home | | | |
|---|---|---|---|---|---|---|---|---|---|---|---|---|---|---|---|---|
| | DF1 | DF2 | FStat | p | DF1 | DF2 | FStat | p | DF1 | DF2 | FStat | p | DF1 | DF2 | FStat | p |
| Light (log lux) | 5 | 3153075 | 83912 | <0.0001 | 23 | 3153075 | 186037 | <0.0001 | 3 | 3153075 | 14562 | <0.0001 | 69 | 3153075 | 3148 | <0.0001 |
| Temperature ($^{o}$C) | 5 | 3144684 | 227267 | <0.0001 | 23 | 3144684 | 7360 | <0.0001 | 3 | 3144684 | 2017 | <0.0001 | 69 | 3144684 | 38033 | <0.0001 |

Table S2: **Daily and seasonal variation of the light and temperature environment.** Results of a linear fixed effect model for average hourly light (log lux) and temperature (°C) for the fixed effects of hour of the day, two-month period, room and home. Hour, two-month period, room and home were all entered as categorical variables.

| | Two-month period | | | |
|---|---|---|---|---|
| | DF1 | DF2 | FStat | p value |
| Bright light, lounge (hours > 500 lux) | 5 | 26517 | 1406 | <0.0001 |
| Geometric mean daily light, lounge (lux) | 5 | 26517 | 2034 | <0.0001 |
| Mean nocturnal temperature, bedroom ($^{o}$C) | 5 | 26517 | 4377 | <0.0001 |
| Temperature amplitude, bedroom ($^{o}$C) | 5 | 26494 | 764 | <0.0001 |

Table S3: **Seasonal variation of the light and temperature environment.** Results of a linear mixed effect model for various *d*aily indoor light and temperature measures with the fixed effect of two-month period and the random effect of home.

| | Number of observations | Mean | Standard deviation | Standard error | 95th confidence interval (low) | 95th confidence interval (high) | Minimum | Maximum | Median | 25th percentile | 75th percentile | IQR | Intraclass correlation coefficient, ICC |
|---|---|---|---|---|---|---|---|---|---|---|---|---|---|
| TOTAL TIME IN-BED PER 24 HOURS | | | | | | | | | | | | | |
| Total time in-bed (h:mm) | 70 | 8:48 | 2:05 | 0:15 | 8:18 | 9:18 | 3:11 | 14:36 | 8:48 | 7:45 | 10:01 | 2:16 | 0.52 |
| TIME IN-BED INITIATED DURING THE DAY | | | | | | | | | | | | | |
| NUMBER & DURATION | | | | | | | | | | | | | |
| Number of in-bed periods initiated during the day (per day) | 70 | 0.14 | 0.24 | 0.03 | 0.08 | 0.20 | 0.00 | 1.34 | 0.04 | 0.01 | 0.12 | 0.11 | 0.33 |
| Total duration of in-bed periods initiated during the day (h:mm) | 66 | 1:39 | 0:57 | 0:07 | 1:24 | 1:53 | 0:15 | 5:19 | 1:18 | 0:60 | 2:09 | 1:10 | 0.21 |
| TIMING | | | | | | | | | | | | | |
| Onset time of the longest in-bed period initiated during the day (hh:mm) | 66 | 12:24 | 2:13 | 0:16 | 11:51 | 12:56 | 07:16 | 16:21 | 12:40 | 11:04 | 14:09 | 3:06 | 0.34 |
| TIME IN-BED INITIATED DURING THE NIGHT | | | | | | | | | | | | | |
| DURATION | | | | | | | | | | | | | |
| Duration of the major nocturnal in-bed period (h:mm) | 70 | 8:47 | 2:09 | 0:15 | 8:17 | 9:18 | 3:11 | 14:39 | 8:54 | 7:36 | 9:42 | 2:06 | 0.51 |
| TIMING | | | | | | | | | | | | | |
| Onset time of the major nocturnal in-bed period (hh:mm) | 70 | 23:05 | 1:18 | 0:09 | 22:47 | 23:24 | 19:07 | 02:04 | 23:10 | 22:17 | 23:49 | 1:32 | 0.41 |
| Onset time of the major nocturnal in-bed period (GMT) (hh:mm) | 70 | 22:31 | 1:17 | 0:09 | 22:13 | 22:50 | 18:35 | 01:30 | 22:36 | 21:44 | 23:13 | 1:29 | 0.39 |
| Time from sunset to the onset of the major nocturnal in-bed period (h:mm) | 70 | 4:28 | 1:19 | 0:09 | 4:09 | 4:47 | 0:28 | 7:36 | 4:25 | 3:45 | 5:12 | 1:27 | 0.22 |
| Offset time of the major nocturnal in-bed period (hh:mm) | 70 | 07:51 | 1:33 | 0:11 | 07:28 | 08:13 | 03:06 | 11:36 | 08:05 | 06:59 | 08:54 | 1:56 | 0.40 |
| Offset time of the major nocturnal in-bed period (GMT) (hh:mm) | 70 | 07:17 | 1:33 | 0:11 | 06:54 | 07:39 | 02:23 | 11:03 | 07:29 | 06:25 | 08:17 | 1:52 | 0.38 |
| Time from sunrise to the offset of the major nocturnal in-bed period (h:mm) | 70 | 1:19 | 1:35 | 0:11 | 0:56 | 1:42 | -3:30 | 5:01 | 1:31 | 0:19 | 2:25 | 2:07 | 0.35 |
| FRAGMENTATION | | | | | | | | | | | | | |
| Number of bed exits (per major nocturnal in-bed period) | 70 | 3.10 | 2.65 | 0.32 | 2.47 | 3.73 | 0.11 | 17.63 | 2.27 | 1.52 | 4.01 | 2.49 | 0.32 |
| Bed exit rate (exits/h) | 70 | 0.40 | 0.37 | 0.04 | 0.31 | 0.48 | 0.01 | 1.71 | 0.27 | 0.18 | 0.50 | 0.32 | 0.29 |
| Median duration of bed exits (h:mm) | 70 | 0:06 | 0:04 | 0:00 | 0:05 | 0:07 | 0:01 | 0:27 | 0:05 | 0:04 | 0:07 | 0:03 | 0.29 |
| Wake after sleep onset (h:mm) | 70 | 1:15 | 0:41 | 0:05 | 1:05 | 1:25 | 0:10 | 3:48 | 1:10 | 0:48 | 1:32 | 0:45 | 0.30 |
| Median duration of wake bouts (h:mm) | 70 | 0:15 | 0:08 | 0:01 | 0:13 | 0:17 | 0:05 | 0:39 | 0:14 | 0:09 | 0:19 | 0:09 | 0.15 |
| Median duration of sleep bouts (h:mm) | 70 | 1:38 | 0:57 | 0:07 | 1:24 | 1:51 | 0:24 | 4:42 | 1:21 | 1:02 | 2:01 | 0:59 | 0.33 |
| Sleep efficiency (%) | 70 | 75.69 | 13.60 | 1.63 | 72.45 | 78.93 | 22.83 | 95.78 | 77.81 | 71.52 | 85.49 | 13.98 | 0.51 |
| PHYSIOLOGY | | | | | | | | | | | | | |
| MEAN & DURATION | | | | | | | | | | | | | |
| Mean respiratory rate (breaths/min) | 70 | 14.89 | 2.16 | 0.26 | 14.37 | 15.40 | 11.42 | 20.41 | 14.52 | 13.31 | 15.98 | 2.67 | 0.85 |
| Snoring duration (h:mm) | 70 | 0:17 | 0:21 | 0:02 | 0:12 | 0:22 | 0:00 | 1:19 | 0:08 | 0:03 | 0:24 | 0:21 | 0.41 |
| Mean heart rate (beats/min) | 70 | 59.47 | 6.69 | 0.80 | 57.88 | 61.07 | 47.09 | 75.78 | 58.93 | 54.26 | 64.25 | 9.99 | 0.80 |
| Std dev. heart rate (beats/min) | 70 | 3.97 | 1.07 | 0.13 | 3.71 | 4.22 | 0.91 | 6.58 | 3.80 | 3.39 | 4.62 | 1.22 | 0.44 |
| TIMING | | | | | | | | | | | | | |
| Time of minimum heart rate (hh:mm) | 70 | 04:36 | 1:30 | 0:11 | 04:15 | 04:58 | 00:36 | 07:20 | 04:40 | 03:38 | 05:50 | 2:12 | 0.24 |
| Time of minimum heart rate (GMT) (hh:mm) | 70 | 04:03 | 1:31 | 0:11 | 03:41 | 04:24 | 00:05 | 06:48 | 04:04 | 03:05 | 05:10 | 2:05 | 0.23 |
| Time from heart rate minimum to nocturnal in-bed period offset (h:mm) | 70 | 3:17 | 1:18 | 0:09 | 2:58 | 3:36 | 1:21 | 8:43 | 3:05 | 2:23 | 3:56 | 1:34 | 0.21 |
| ENVIRONMENT | | | | | | | | | | | | | |
| Geometric mean daily light, lounge (lux) | 70 | 46.31 | 25.14 | 3.01 | 40.32 | 52.31 | 17.18 | 155.65 | 40.12 | 30.80 | 58.02 | 27.22 | 0.33 |
| Hours bright light, lounge (h:mm > 500 lux) | 70 | 2.60 | 2.33 | 0.28 | 2.04 | 3.15 | 0.00 | 9.56 | 1.86 | 0.92 | 4.02 | 3.10 | 0.44 |
| Mean nocturnal temperature, bedroom ($^{o}$C) | 70 | 21.05 | 2.20 | 0.26 | 20.53 | 21.57 | 16.28 | 25.84 | 21.00 | 19.48 | 22.21 | 2.73 | 0.47 |
| Amplitude temperature, bedroom ($^{o}$C) | 70 | 2.76 | 1.14 | 0.14 | 2.49 | 3.03 | 1.07 | 5.86 | 2.62 | 1.91 | 3.22 | 1.31 | 0.40 |

Table S4: **Descriptive statistics for the measures of sleep behaviour and physiology and the light and temperature environment** Descriptive statistics describing the distribution of the $n = 70$ participant mean values for the 25 different measures of sleep behaviour and physiology and 4 measures of the light and temperature environment. The intraclass correlation coefficient for each measure is also included. For all measures, the reported mean is the mean of the participant mean values, i.e. from the approximately 26,000 days of data, the individual participant mean values were first calculated. Then the mean of the participant values was found.

| | Initial age | | Baseline MMSE | | Baseline NPI (total) | | Baseline NPI (sleep) | | Geometric mean daily light, lounge (lux) | | Hours bright light, lounge (h:mm > 500 lux) | | Mean nocturnal temperature, bedroom (^oC) | | Amplitude temperature, bedroom ("oC) | |
|---|---|---|---|---|---|---|---|---|---|---|---|---|---|---|---|---|
| | R | p | R | p | R | p | R | p | R | p | R | p | R | p | R | p |
| TOTAL TIME IN-BED PER 24 HOURS | | | | | | | | | | | | | | | | |
| Total time in-bed (h:mm) | 0.055 | 0.6504 | -0.237 | 0.0537 | 0.110 | 0.3745 | -0.258 | 0.0350 | -0.109 | 0.3703 | -0.114 | 0.3480 | 0.036 | 0.7656 | 0.030 | 0.8071 |
| TIME IN-BED INITIATED DURING THE DAY | | | | | | | | | | | | | | | | |
| NUMBER & DURATION | | | | | | | | | | | | | | | | |
| Number of in-bed periods initiated during the day (per day) | -0.317 | 0.0074 | 0.172 | 0.1652 | 0.044 | 0.7256 | 0.157 | 0.2053 | -0.182 | 0.1318 | -0.178 | 0.1394 | 0.278 | 0.0199 | -0.260 | 0.0298 |
| Total duration of in-bed periods initiated during the day (h:mm) | -0.001 | 0.9922 | -0.024 | 0.8510 | 0.060 | 0.6426 | 0.115 | 0.3696 | 0.013 | 0.9161 | -0.033 | 0.7906 | 0.352 | 0.0037 | -0.334 | 0.0061 |
| TIMING | | | | | | | | | | | | | | | | |
| Onset time of the longest in-bed period initiated during the day (hh:mm) | -0.245 | 0.0473 | 0.382 | 0.0020 | -0.022 | 0.8642 | -0.096 | 0.4556 | 0.060 | 0.6350 | 0.003 | 0.9785 | 0.008 | 0.9473 | 0.003 | 0.9792 |
| TIME IN-BED INITIATED DURING THE NIGHT | | | | | | | | | | | | | | | | |
| DURATION | | | | | | | | | | | | | | | | |
| Duration of the major nocturnal in-bed period (h:mm) | 0.059 | 0.6270 | -0.264 | 0.0310 | 0.028 | 0.8240 | -0.299 | 0.0139 | -0.058 | 0.6306 | -0.059 | 0.6260 | -0.100 | 0.4078 | 0.110 | 0.3651 |
| TIMING | | | | | | | | | | | | | | | | |
| Onset time of the major nocturnal in-bed period (hh:mm) | -0.151 | 0.2136 | 0.352 | 0.0035 | -0.071 | 0.5683 | 0.268 | 0.0281 | 0.007 | 0.9559 | -0.041 | 0.7344 | -0.096 | 0.4263 | -0.063 | 0.6050 |
| Onset time of the major nocturnal in-bed period (GMT) (hh:mm) | -0.159 | 0.1877 | 0.355 | 0.0032 | -0.069 | 0.5788 | 0.275 | 0.0245 | -0.009 | 0.9428 | -0.046 | 0.7053 | -0.098 | 0.4171 | -0.074 | 0.5434 |
| Time from sunset to the onset of the major nocturnal in-bed period (h:mm) | -0.130 | 0.2827 | 0.360 | 0.0028 | -0.065 | 0.5985 | 0.265 | 0.0305 | -0.083 | 0.4928 | -0.097 | 0.4237 | -0.133 | 0.2702 | -0.092 | 0.4487 |
| Offset time of the major nocturnal in-bed period (hh:mm) | -0.063 | 0.6066 | -0.083 | 0.5060 | 0.072 | 0.5634 | -0.157 | 0.2056 | -0.077 | 0.5231 | -0.138 | 0.2547 | -0.114 | 0.3475 | 0.129 | 0.2858 |
| Offset time of the major nocturnal in-bed period (GMT) (hh:mm) | -0.054 | 0.6584 | -0.089 | 0.4732 | 0.076 | 0.5389 | -0.149 | 0.2300 | -0.087 | 0.4726 | -0.145 | 0.2326 | -0.127 | 0.2944 | 0.118 | 0.3316 |
| Time from sunrise to the offset of the major nocturnal in-bed period (h:mm) | -0.054 | 0.6587 | -0.083 | 0.5038 | 0.101 | 0.4169 | -0.135 | 0.2755 | -0.044 | 0.7166 | -0.124 | 0.3082 | -0.101 | 0.4052 | 0.142 | 0.2419 |
| FRAGMENTATION | | | | | | | | | | | | | | | | |
| Number of bed exits (per major nocturnal in-bed period) | -0.020 | 0.8702 | -0.012 | 0.9242 | 0.128 | 0.3025 | 0.208 | 0.0908 | -0.057 | 0.6380 | 0.010 | 0.9334 | -0.138 | 0.2556 | 0.076 | 0.5283 |
| Bed exit rate (exits/h) | -0.021 | 0.8610 | 0.016 | 0.8977 | 0.124 | 0.3173 | 0.271 | 0.0266 | -0.039 | 0.7483 | 0.024 | 0.8412 | -0.097 | 0.4249 | 0.039 | 0.7450 |
| Median duration of bed exits (h:mm) | 0.127 | 0.2934 | 0.092 | 0.4612 | -0.012 | 0.9218 | 0.104 | 0.4019 | -0.008 | 0.9494 | 0.013 | 0.9145 | 0.409 | 0.0005 | -0.210 | 0.0805 |
| Wake after sleep onset (h:mm) | 0.101 | 0.4050 | -0.133 | 0.2846 | 0.224 | 0.0685 | 0.159 | 0.1993 | -0.212 | 0.0782 | -0.224 | 0.0625 | 0.282 | 0.0181 | -0.122 | 0.3122 |
| Median duration of wake bouts (h:mm) | 0.119 | 0.3279 | 0.107 | 0.3869 | 0.073 | 0.5546 | 0.142 | 0.2510 | -0.135 | 0.2659 | -0.173 | 0.1525 | 0.388 | 0.0010 | -0.239 | 0.0462 |
| Median duration of sleep bouts (h:mm) | -0.008 | 0.9460 | -0.025 | 0.8385 | -0.155 | 0.2095 | -0.370 | 0.0021 | 0.003 | 0.9779 | 0.042 | 0.7297 | 0.071 | 0.5567 | -0.068 | 0.5772 |
| Sleep efficiency (%) | -0.180 | 0.1368 | 0.004 | 0.9768 | -0.179 | 0.1477 | -0.325 | 0.0073 | 0.183 | 0.1294 | 0.178 | 0.1410 | -0.266 | 0.0265 | 0.181 | 0.1340 |
| NOCTURNAL PHYSIOLOGY | | | | | | | | | | | | | | | | |
| MEAN & DURATION | | | | | | | | | | | | | | | | |
| Mean respiratory rate (breaths/min) | 0.151 | 0.2108 | 0.093 | 0.4544 | -0.098 | 0.4317 | -0.111 | 0.3723 | 0.082 | 0.5005 | 0.069 | 0.5693 | 0.101 | 0.4063 | 0.087 | 0.4710 |
| Snoring duration (h:mm) | -0.163 | 0.1782 | -0.058 | 0.6424 | 0.010 | 0.9343 | 0.122 | 0.3248 | 0.260 | 0.0294 | 0.348 | 0.0032 | 0.081 | 0.5043 | 0.022 | 0.8546 |
| Mean heart rate (beats/min) | 0.331 | 0.0051 | -0.093 | 0.4557 | -0.019 | 0.8774 | 0.095 | 0.4434 | -0.057 | 0.6380 | -0.027 | 0.8247 | 0.162 | 0.1812 | 0.049 | 0.6881 |
| Std dev. heart rate (beats/min) | -0.006 | 0.9639 | -0.092 | 0.4572 | -0.161 | 0.1919 | -0.071 | 0.5668 | -0.126 | 0.2966 | -0.107 | 0.3767 | -0.186 | 0.1231 | 0.183 | 0.1297 |
| TIMING | | | | | | | | | | | | | | | | |
| Time of minimum heart rate (hh:mm) | -0.159 | 0.1896 | 0.235 | 0.0553 | -0.108 | 0.3850 | -0.037 | 0.7666 | -0.117 | 0.3341 | -0.150 | 0.2144 | -0.093 | 0.4432 | 0.171 | 0.1562 |
| Time of minimum heart rate (GMT) (hh:mm) | -0.153 | 0.2055 | 0.241 | 0.0498 | -0.105 | 0.3969 | -0.034 | 0.7828 | -0.131 | 0.2799 | -0.157 | 0.1945 | -0.099 | 0.4146 | 0.177 | 0.1420 |
| Time from heart rate minimum to nocturnal in-bed period offset (h:mm) | 0.072 | 0.5561 | -0.355 | 0.0032 | 0.227 | 0.0645 | -0.207 | 0.0929 | -0.018 | 0.8813 | -0.064 | 0.6006 | -0.043 | 0.7202 | -0.105 | 0.3844 |

Table S5: **Correlation between bedmat variables and measures of age, cognition, behaviour and environmental light and temperature.** Correlation coefficients (Spearman's rho) and p values were calculated as the correlation between participant mean values (n=70).

| | | Two-month period | | | Gender | | | | Alzheimer's diagnosis | | | | Initial age | | | |
|---|---|---|---|---|---|---|---|---|---|---|---|---|---|---|---|---|
| | Intercept | DF | Fstat | p | DF | Fstat | p | Man effect | DF | Fstat | p | Not Alzheimer's effect | DF | Fstat | p | Initial age effect (per y) |
| TOTAL TIME IN-BED PER 24 HOURS | | | | | | | | | | | | | | | | |
| Total time in-bed (h) | 8.41 | 26138 | 34.36 | 4.0E-35 | 26138 | 0.11 | 0.7412 | 0.18 | 26138 | 1.98 | 0.1593 | -0.79 | 26138 | 0.03 | 0.8681 | 0.005 |
| TIME IN-BED INITIATED DURING THE DAY | | | | | | | | | | | | | | | | |
| NUMBER & DURATION | | | | | | | | | | | | | | | | |
| Number of in-bed periods initiated during the day (per day) | 0.42 | 26138 | 4.69 | 2.8E-04 | 26138 | 0.13 | 0.7231 | 0.02 | 26138 | 0.21 | 0.6499 | 0.03 | 26138 | 1.14 | 0.2849 | -0.004 |
| Total duration of in-bed periods initiated during the day (h) | 1.13 | 3276 | 0.77 | 5.7E-01 | 3276 | 0.00 | 0.9631 | -0.01 | 3276 | 0.35 | 0.5540 | -0.15 | 3276 | 0.40 | 0.5270 | 0.008 |
| TIMING | | | | | | | | | | | | | | | | |
| Onset time of the longest in-bed period initiated during the day (h) | 16.23 | 3276 | 1.53 | 1.8E-01 | 3276 | 3.77 | 0.0522 | 0.99 | 3276 | 0.54 | 0.4642 | -0.39 | 3276 | 4.00 | 0.0455 | -0.055 |
| TIME IN-BED INITIATED DURING THE NIGHT | | | | | | | | | | | | | | | | |
| DURATION | | | | | | | | | | | | | | | | |
| Duration of the major nocturnal in-bed period (h) | 8.30 | 26138 | 29.89 | 2.1E-30 | 26138 | 0.01 | 0.9174 | -0.06 | 26138 | 2.05 | 0.1523 | -0.82 | 26138 | 0.07 | 0.7940 | 0.008 |
| TIMING | | | | | | | | | | | | | | | | |
| Onset time of the major nocturnal in-bed period (h) | 0.84 | 26138 | 1.17 | 3.2E-01 | 26138 | 0.23 | 0.6336 | 0.16 | 26138 | 0.36 | 0.5494 | 0.21 | 26138 | 1.64 | 0.2002 | -0.024 |
| Onset time of the major nocturnal in-bed period (GMT) (h) | -0.14 | 26138 | 402.36 | 0.0E+00 | 26138 | 0.24 | 0.6262 | 0.16 | 26138 | 0.34 | 0.5614 | 0.20 | 26138 | 1.62 | 0.2031 | -0.023 |
| Time from sunset to the onset of the major nocturnal in-bed period (h) | 6.24 | 26138 | 6469.65 | 0.0E+00 | 26138 | 0.19 | 0.6669 | 0.14 | 26138 | 0.33 | 0.5645 | 0.20 | 26138 | 1.69 | 0.1933 | -0.024 |
| Offset time of the major nocturnal in-bed period (hh:mm) | 8.85 | 26138 | 24.76 | 5.7E-25 | 26138 | 0.12 | 0.7334 | 0.13 | 26138 | 2.28 | 0.1313 | -0.63 | 26138 | 0.34 | 0.5592 | -0.013 |
| Offset time of the major nocturnal in-bed period (GMT) (h) | 7.87 | 26138 | 450.52 | 0.0E+00 | 26138 | 0.12 | 0.7280 | 0.14 | 26138 | 2.32 | 0.1277 | -0.63 | 26138 | 0.33 | 0.5662 | -0.013 |
| Time from sunrise to the offset of the major nocturnal in-bed period (h) | 1.81 | 26138 | 1252.28 | 0.0E+00 | 26138 | 0.14 | 0.7034 | 0.15 | 26138 | 2.28 | 0.1311 | -0.63 | 26138 | 0.32 | 0.5738 | -0.012 |
| FRAGMENTATION | | | | | | | | | | | | | | | | |
| Number of bed exits (per major nocturnal in-bed period) | 6.37 | 26138 | 48.87 | 1.6E-50 | 26138 | 5.67 | 0.0173 | -1.55 | 26138 | 0.02 | 0.8843 | -0.10 | 26138 | 0.27 | 0.6035 | -0.019 |
| Bed exit rate (exits/h) | 0.56 | 26138 | 40.48 | 1.3E-41 | 26138 | 8.14 | 0.0043 | -0.25 | 26138 | 1.01 | 0.3160 | 0.09 | 26138 | 0.03 | 0.8626 | 0.001 |
| Median duration of bed exits (h) | 0.04 | 26138 | 9.16 | 1.0E-08 | 26138 | 0.04 | 0.8379 | 0.00 | 26138 | 3.21 | 0.0733 | 0.03 | 26138 | 0.38 | 0.5352 | 0.001 |
| Wake after sleep onset (h) | 0.43 | 26138 | 3.77 | 2.1E-03 | 26138 | 0.65 | 0.4204 | -0.14 | 26138 | 0.00 | 0.9599 | -0.01 | 26138 | 1.41 | 0.2352 | 0.012 |
| Median duration of wake bouts (h) | 0.11 | 26138 | 3.35 | 5.1E-03 | 26138 | 0.52 | 0.4693 | 0.02 | 26138 | 3.24 | 0.0720 | 0.06 | 26138 | 0.51 | 0.4747 | 0.001 |
| Median duration of sleep bouts (h) | 0.55 | 26138 | 5.86 | 2.1E-05 | 26138 | 0.60 | 0.4372 | 0.19 | 26138 | 0.04 | 0.8485 | -0.05 | 26138 | 0.72 | 0.3958 | 0.012 |
| Sleep efficiency (%) | 97.81 | 26138 | 4.00 | 1.3E-03 | 26138 | 1.09 | 0.2956 | 3.54 | 26138 | 2.04 | 0.1530 | -5.12 | 26138 | 2.30 | 0.1297 | -0.289 |
| NOCTURNAL PHYSIOLOGY | | | | | | | | | | | | | | | | |
| MEAN & DURATION | | | | | | | | | | | | | | | | |
| Mean respiratory rate (breaths/min) | 11.71 | 25801 | 181.86 | 6.1E-191 | 25801 | 0.12 | 0.7269 | 0.19 | 25801 | 0.62 | 0.4317 | 0.46 | 25801 | 1.40 | 0.2363 | 0.036 |
| Snoring duration (h) | 1.03 | 26138 | 7.57 | 4.1E-07 | 26138 | 0.65 | 0.4201 | 0.07 | 26138 | 0.65 | 0.4191 | -0.07 | 26138 | 3.94 | 0.0471 | -0.010 |
| Mean heart rate (beats/min) | 41.87 | 25907 | 25.76 | 5.0E-26 | 25907 | 0.31 | 0.5801 | -0.90 | 25907 | 0.03 | 0.8718 | -0.28 | 25907 | 6.09 | 0.0136 | 0.227 |
| Std dev. heart rate (beats/min) | 4.67 | 25907 | 5.73 | 2.7E-05 | 25907 | 5.22 | 0.0223 | -0.61 | 25907 | 0.42 | 0.5165 | 0.18 | 25907 | 0.14 | 0.7082 | -0.006 |
| TIMING | | | | | | | | | | | | | | | | |
| Time of minimum heart rate (h) | 6.43 | 25907 | 10.28 | 7.3E-10 | 25907 | 0.04 | 0.8440 | -0.08 | 25907 | 0.24 | 0.6242 | 0.20 | 25907 | 1.33 | 0.2496 | -0.025 |
| Time of minimum heart rate (GMT) (h) | 5.44 | 25907 | 194.39 | 4.9E-204 | 25907 | 0.04 | 0.8494 | -0.07 | 25907 | 0.22 | 0.6356 | 0.19 | 25907 | 1.30 | 0.2539 | -0.025 |
| Time from heart rate minimum to nocturnal in-bed period offset (h) | 2.67 | 25907 | 12.38 | 5.1E-12 | 25907 | 0.27 | 0.6027 | 0.17 | 25907 | 5.70 | 0.0169 | -0.81 | 25907 | 0.29 | 0.5917 | 0.010 |

Table S6: **Seasonal effects on the duration, timing and fragmentation of bed occupancy and physiology.** Results from a linear mixed effect model for the seasonal variation of the 25 measures of bed occupancy, physiology and a marker of the biological clock controlling for gender, age and diagnosis. One of the participants did not have a diagnosis, so only 69 of the 70 participants were included in this analysis.

| | | Time-in-study | | | | Hours of bright light (>500 lux) | | | | Mean nocturnal temperature | | | | Temperature amplitude | | | |
|---|---|---|---|---|---|---|---|---|---|---|---|---|---|---|---|---|---|
| | Intercept | DF | Fstat | p | Time-in-study effect (per year) | DF | Fstat | p | Hours of bright light effect (per h) | DF | Fstat | p | Nocturnal temperature effect (per C°) | DF | Fstat | p | Temperature amplitude effect (per C°) |
| TOTAL TIME IN-BED PER 24 HOURS | | | | | | | | | | | | | | | | | |
| Total time in-bed (h) | 8.43 | 26111 | 2.07 | 1.50E-01 | 0.0428 | 26111 | 12.30 | 4.54E-04 | -0.019 | 26111 | 0.41 | 5.23E-01 | -0.005 | 26111 | 90.80 | 1.72E-21 | 0.0925 |
| TIME IN-BED INITIATED DURING THE DAY | | | | | | | | | | | | | | | | | |
| NUMBER & DURATION | | | | | | | | | | | | | | | | | |
| Number of in-bed periods initiated during the day (per day) | 0.34 | 26111 | 34.62 | 4.06E-09 | 0.0295 | 26111 | 0.68 | 4.10E-01 | 0.001 | 26111 | 7.26 | 7.04E-03 | 0.003 | 26111 | 0.12 | 7.32E-01 | 0.0006 |
| Total duration of in-bed periods initiated during the day (h) | 1.44 | 3265 | 36.23 | 1.95E-09 | 0.4125 | 3265 | 0.49 | 4.83E-01 | -0.008 | 3265 | 1.50 | 2.21E-01 | -0.019 | 3265 | 2.97 | 8.51E-02 | 0.0408 |
| TIMING | | | | | | | | | | | | | | | | | |
| Onset time of the longest in-bed period initiated during the day (h) | 16.52 | 3265 | 5.41 | 2.01E-02 | -0.2696 | 3265 | 1.18 | 2.77E-01 | -0.021 | 3265 | 0.18 | 6.67E-01 | -0.012 | 3265 | 0.38 | 5.36E-01 | 0.0249 |
| TIME IN-BED INITIATED DURING THE NIGHT | | | | | | | | | | | | | | | | | |
| DURATION | | | | | | | | | | | | | | | | | |
| Duration of the major nocturnal in-bed period (h) | 8.59 | 26111 | 10.06 | 1.52E-03 | -0.1004 | 26111 | 10.47 | 1.22E-03 | -0.018 | 26111 | 3.65 | 5.62E-02 | -0.015 | 26111 | 71.41 | 3.06E-17 | 0.0873 |
| TIMING | | | | | | | | | | | | | | | | | |
| Onset time of the major nocturnal in-bed period (h) | 1.80 | 26111 | 42.23 | 8.28E-11 | -0.1507 | 26111 | 3.48 | 6.22E-02 | 0.008 | 26111 | 64.18 | 1.18E-15 | -0.045 | 26111 | 1.60 | 2.05E-01 | -0.0096 |
| Onset time of the major nocturnal in-bed period (GMT) (h) | 0.99 | 26111 | 55.05 | 1.21E-13 | -0.1736 | 26111 | 0.36 | 5.51E-01 | -0.003 | 26111 | 86.02 | 1.91E-20 | -0.053 | 26111 | 0.04 | 8.38E-01 | 0.0016 |
| Time from sunset to the onset of the major nocturnal in-bed period (h) | 8.58 | 26111 | 44.30 | 2.87E-11 | -0.1639 | 26111 | 61.19 | 5.37E-15 | -0.035 | 26111 | 339.60 | 2.35E-75 | -0.110 | 26111 | 16.21 | 5.68E-05 | 0.0323 |
| Offset time of the major nocturnal in-bed period (hh:mm) | 10.11 | 26111 | 55.96 | 7.64E-14 | -0.2137 | 26111 | 2.95 | 8.58E-02 | -0.009 | 26111 | 78.44 | 8.78E-19 | -0.061 | 26111 | 75.38 | 4.10E-18 | 0.0809 |
| Offset time of the major nocturnal in-bed period (GMT) (h) | 9.30 | 26111 | 67.87 | 1.83E-16 | -0.2366 | 26111 | 13.70 | 2.15E-04 | -0.019 | 26111 | 97.92 | 4.78E-23 | -0.069 | 26111 | 96.53 | 9.63E-23 | 0.0920 |
| Time from sunrise to the offset of the major nocturnal in-bed period (h) | 2.19 | 26111 | 62.85 | 2.32E-15 | -0.2299 | 26111 | 10.18 | 1.42E-03 | 0.017 | 26111 | 7.97 | 4.77E-03 | -0.020 | 26111 | 36.27 | 1.74E-09 | 0.0570 |
| FRAGMENTATION | | | | | | | | | | | | | | | | | |
| Number of bed exits (per major nocturnal in-bed period) | 7.72 | 26111 | 81.46 | 1.91E-19 | -0.5166 | 26111 | 13.16 | 2.87E-04 | -0.037 | 26111 | 8.98 | 2.73E-03 | -0.042 | 26111 | 70.57 | 4.67E-17 | -0.1568 |
| Bed exit rate (exits/h) | 0.73 | 26111 | 29.99 | 4.38E-08 | -0.0470 | 26111 | 4.77 | 2.89E-02 | -0.003 | 26111 | 7.33 | 6.78E-03 | -0.006 | 26111 | 67.37 | 2.35E-16 | -0.0230 |
| Median duration of bed exits (h) | 0.01 | 26111 | 0.63 | 4.28E-01 | 0.0013 | 26111 | 5.23 | 2.22E-02 | -0.001 | 26111 | 11.17 | 8.33E-04 | 0.001 | 26111 | 0.06 | 8.08E-01 | -0.0001 |
| Wake after sleep onset (h) | 0.20 | 26111 | 8.88 | 2.88E-03 | -0.0469 | 26111 | 7.98 | 4.74E-03 | -0.008 | 26111 | 13.58 | 2.29E-04 | 0.014 | 26111 | 1.42 | 2.34E-01 | -0.0061 |
| Median duration of wake bouts (h) | 0.01 | 26111 | 14.40 | 1.48E-04 | 0.0171 | 26111 | 0.76 | 3.85E-01 | -0.001 | 26111 | 18.67 | 1.56E-05 | 0.005 | 26111 | 0.74 | 3.90E-01 | 0.0013 |
| Median duration of sleep bouts (h) | 0.18 | 26111 | 199.99 | 3.09E-45 | 0.2796 | 26111 | 0.99 | 3.20E-01 | 0.004 | 26111 | 6.18 | 1.30E-02 | 0.012 | 26111 | 0.49 | 4.84E-01 | -0.0045 |
| Sleep efficiency (%) | 100.25 | 26111 | 5.75 | 1.65E-02 | -0.4776 | 26111 | 0.50 | 4.78E-01 | -0.025 | 26111 | 6.04 | 1.40E-02 | -0.119 | 26111 | 8.02 | 4.63E-03 | 0.1841 |
| NOCTURNAL PHYSIOLOGY | | | | | | | | | | | | | | | | | |
| MEAN & DURATION | | | | | | | | | | | | | | | | | |
| Mean respiratory rate (breaths/min) | 9.05 | 25774 | 23.84 | 1.05E-06 | 0.0635 | 25774 | 17.89 | 2.34E-05 | -0.010 | 25774 | 1753.08 | 0.00E+00 | 0.134 | 25774 | 4.63 | 3.14E-02 | 0.0092 |
| Snoring duration (h) | 1.27 | 26111 | 142.93 | 7.40E-33 | -0.0733 | 26111 | 8.63 | 3.32E-03 | 0.003 | 26111 | 56.64 | 5.39E-14 | -0.011 | 26111 | 0.10 | 7.55E-01 | -0.0006 |
| Mean heart rate (beats/min) | 40.39 | 25880 | 4.16 | 4.14E-02 | -0.1006 | 25880 | 0.12 | 7.26E-01 | 0.003 | 25880 | 36.33 | 1.69E-09 | 0.073 | 25880 | 12.90 | 3.29E-04 | 0.0581 |
| Std dev. heart rate (beats/min) | 4.78 | 25880 | 5.95 | 1.47E-02 | -0.0443 | 25880 | 5.92 | 1.50E-02 | -0.008 | 25880 | 1.05 | 3.06E-01 | -0.005 | 25880 | 11.08 | 8.72E-04 | 0.0198 |
| TIMING | | | | | | | | | | | | | | | | | |
| Time of minimum heart rate (h) | 7.13 | 25880 | 32.25 | 1.37E-08 | -0.2292 | 25880 | 0.72 | 3.95E-01 | 0.006 | 25880 | 12.69 | 3.68E-04 | -0.035 | 25880 | 33.73 | 6.42E-09 | 0.0766 |
| Time of minimum heart rate (GMT) (h) | 6.32 | 25880 | 38.83 | 4.69E-10 | -0.2520 | 25880 | 0.32 | 5.71E-01 | -0.004 | 25880 | 18.63 | 1.59E-05 | -0.043 | 25880 | 44.16 | 3.09E-11 | 0.0878 |
| Time from heart rate minimum to nocturnal in-bed period offset (h) | 3.19 | 25880 | 0.24 | 6.26E-01 | -0.0181 | 25880 | 6.17 | 1.30E-02 | -0.017 | 25880 | 6.62 | 1.01E-02 | -0.023 | 25880 | 0.02 | 8.77E-01 | -0.0019 |

Table S7: **Effects of light, temperature and time-in-study on the duration, timing and fragmentation of bed occupancy and on physiology.** Linear mixed effect model results for the 25 measures of bed occupancy and physiology, extending the model reported in Table S6 to include environmental variables. Only the results for the additional variables (hours of bright light, mean nocturnal temperature, temperature amplitude and time-in-study) are shown. All other effects are similar to those reported in Supplementary Table S6). One of the participant's did not have a diagnosis, so only 69 of the 70 participants were included in this analysis.

| | Initial age | | Baseline MMSE | | Baseline NPI (total) | | Baseline NPI (sleep) | | Seasonal change in geometric mean daily light, lounge (lux) | | Seasonal change in hours of bright light (>500 lux) | | Seasonal change in mean nocturnal temperature | | Seasonal change in temperature amplitude | | Difference between weekend and week day duration of the major nocturnal in-bed period | | Difference between weekend and week day time of onset of the major nocturnal in-bed period | | Difference between weekend and week day time of offset of the major nocturnal in-bed period | |
|---|---|---|---|---|---|---|---|---|---|---|---|---|---|---|---|---|---|---|---|---|---|---|
| | R | p | R | p | R | p | R | p | R | p | R | p | R | p | R | p | R | p | R | p | R | p |
| Seasonal change in duration of the major nocturnal in-bed period (h) | -0.12 | 0.349 | 0.12 | 0.370 | -0.20 | 0.132 | -0.28 | 0.034 | -0.32 | 0.012 | -0.34 | 0.009 | 0.06 | 0.661 | 0.23 | 0.077 | 0.12 | 0.362 | -0.07 | 0.618 | 0.10 | 0.465 |
| Seasonal change in the onset time of the major nocturnal in-bed period (h) | 0.06 | 0.666 | -0.13 | 0.324 | 0.12 | 0.383 | 0.20 | 0.136 | 0.37 | 0.004 | 0.29 | 0.024 | -0.10 | 0.453 | -0.22 | 0.087 | -0.12 | 0.362 | -0.14 | 0.284 | -0.32 | 0.013 |
| Seasonal change in the offset time of the major nocturnal in-bed period (h) | -0.05 | 0.695 | 0.04 | 0.766 | -0.12 | 0.374 | -0.11 | 0.415 | -0.11 | 0.391 | -0.20 | 0.117 | -0.08 | 0.536 | 0.22 | 0.096 | -0.05 | 0.694 | -0.13 | 0.307 | -0.11 | 0.388 |

Table S8: **Correlation between the seasonal change in duration, onset and offset of the major time-in-bed period with age, MMSE and NPI and the seasonal change in environmental light and temperature.** Correlation coefficients (Spearman's rho) and p values were calculated as the correlation between participant mean values (n=70). The seasonal change for each participant was calculated as the mean winter (Jan-Feb) value minus the mean summer (Jul-Aug) value.

### S5 Seasonal desynchrony

The time of the heart rate minimum is a hypothesised marker for the timing of the biological clock with heart rate tending to drop overnight to a minimum and then rising before wake. This can be seen in Fig. 1 and is reproduced in Fig. S5 (a) for the “seasonal” participant. An example for one night is shown in Fig. S5 (b). Around January in the transition from Year 2 to Year 3, the time of the heart rate minimum appears to desynchronise from the 24 h day. Light is the major driver of entrainment to the 24 h day and for the timing of the biological clock. For this participant, the environmental sensors in the home indicate that the indoor light environment is very dim, particularly during the winter months, see Fig. S5 (c), well below recommended light levels [1]. To assess whether the low levels of lighting could be a contributing factor to the observed desynchrony we used a mathematical model which has previously been shown to accurately fit individual mean sleep duration and timing using light collected from wearable sensors [2]. We took the first month of environmental light and bedmat sleep timing data (June of Year 1) and, as in [2], tuned two parameters one for sleep duration and one for sleep timing to create a personalised model. We then took the personalised model and fed in the environmental light to predict sleep timing in all subsequent months in two scenarios. First assuming that there was no feedback from the sleep-wake cycle on to the light-dark cycle. Second assuming that light was gated by sleep. Predicted sleep timing is shown for the two cases in (d). Interestingly, we found that when lights were turned on and off according to when measured bedtime occurred, we found that there was a delay in predicted sleep and circadian timing in the winter months. Whereas, when light is gated by predicted sleep-wake timing, desynchrony in the winter occurs. These two scenarios demonstrate the stabiliising effect of imposed lights out / bedtimes on sleep and circadian timing, as highlighted in [3]. One explanation for the difference between the first and seond winter is that a period of ill-health leading to changes in routine, combined with the low levels of ambient light could trigger the observed desynchrony.

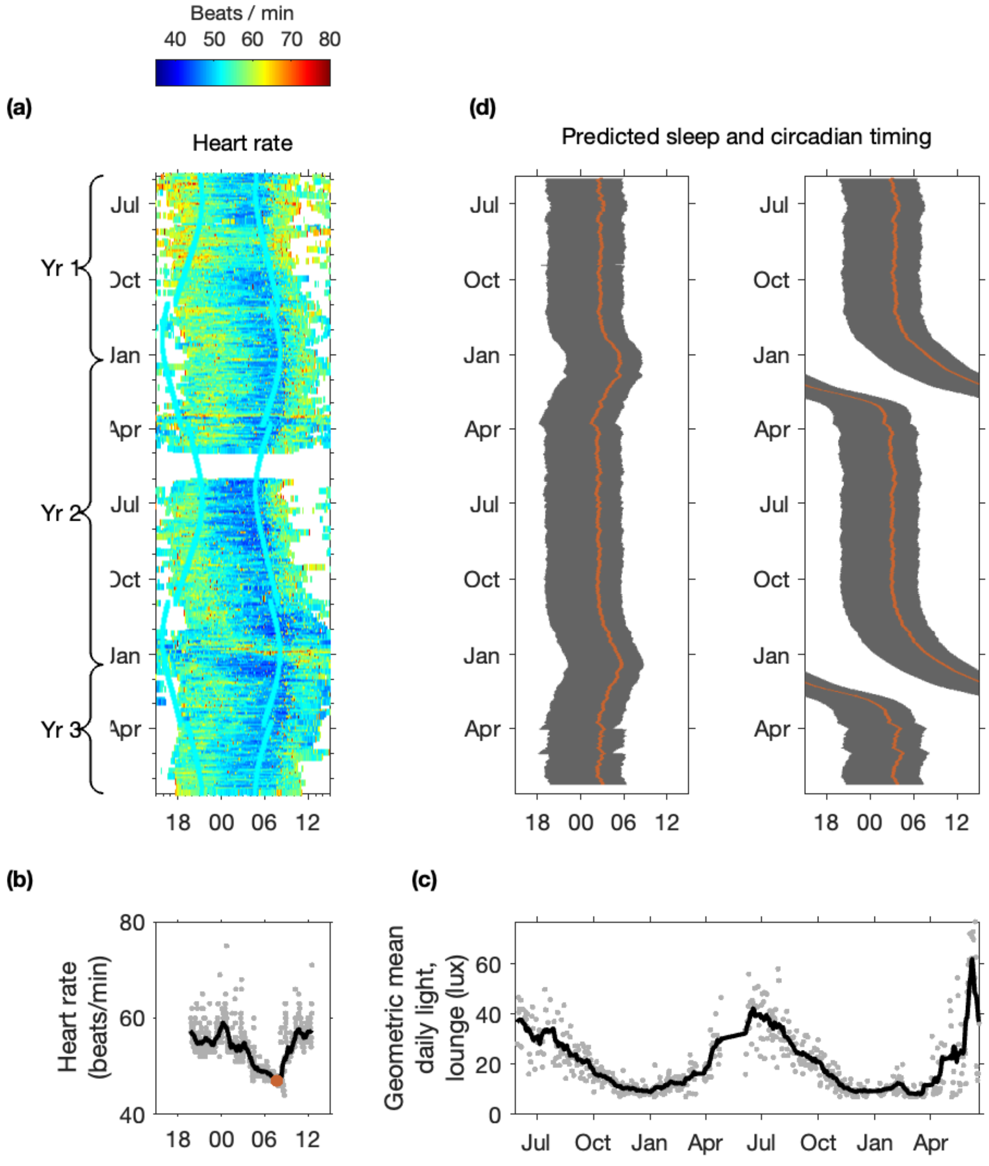

Figure S6: **Desynchrony due to a maladaptive light environment?** a) Raster plot for the heart rate for participant "seasonal", replicated from Fig. 1 (b). Recording of heart rate during sleep for one night showing how the heart rate dips to a minimum. Superimposed on the raw data is a 60 minute moving average, with the circular marker indicating the minimum. (c) Mean daily light levels in the lounge for participant "seasonal". (d) Raster plots of sleep predictions. In the left hand panel, it is assumed that light is not gated by sleep. In the right hand panel, it is assumed that the measured light is gated by sleep.

### Bibliography

[1] Brown T. M., Brainard G. C., Cajochen C., Czeisler C. A., Hanifin J. P., Lockley S.W., and et al. Recommendations for daytime, evening, and nighttime indoor light exposure to best support physiology, sleep, and wakefulness in healthy adults. *PLoS Biol.*, 20:e3001571, 2022.

[2] Skeldon A. C., T. Rodriguez Garcia, S. F. Cleator, C. della Monica, K. K. G. Ravindran, V. L. Revell, and D.-J. Dijk. Method to determine whether sleep phenotypes are driven by endogenous circadian rhythms or environmental light by combining longitudinal data and personalised mathematical models. *PLOS Comput. Biol.*, 19:e1011743, 2023.

[3] A.C. Skeldon, A.J.K. Phillips, and D.-J. Dijk. The effects of self-selected light-dark cycles and social constraints on human sleep and circadian timing: a modeling approach. *Sci Rep*, 7:45158, 2017.
